# The effect of a single session of tDCS on attention in pediatric acquired brain injury: Characterising inter-individual structural and functional network response variability

**DOI:** 10.1101/2025.02.27.25323049

**Authors:** Athena Stein, Kevin A. Caulfield, Mervyn Singh, Justin Riddle, Maximilian A. Friehs, Michael P. Craven, Madeleine J. Groom, Kartik K. Iyer, Karen M. Barlow

## Abstract

**Background:** Approximately 1 in 4 children who sustain an acquired brain injury (ABI) have attention difficulties impacting education, employment, and community participation. These difficulties arise from dysfunction in attention-related brain networks, incentivising the use of transcranial direct current stimulation (tDCS).

**Objective/Hypothesis:** We investigated whether a single tDCS session improved attention following childhood ABI and whether baseline structural connectivity (sc), functional connectivity (fc), attention, and/or simulated electric fields (E-field) explained variability in response.

**Methods:** In a randomised, single-blind, within-subject, sham-controlled trial, 15 children with ABI (mean 12.7 years) and 15 healthy controls (HCs) received three single tDCS sessions (1mA dorsolateral prefrontal cortex [dlPFC], 1mA inferior frontal gyrus [IFG], sham; 20min) during gamified attention training. We examined post-intervention changes in attention according to flanker and stop signal reaction time (RT). We used multi-modal analyses (high-density electroencephalography [HD-EEG], diffusion tensor imaging, magnetic resonance imaging) to investigate inter-individual variability in tDCS response, according to associations between RT change and baseline fc, sc, attention, and E-fields.

**Results:** Although no effect of active versus sham tDCS was found overall, participants with lower theta or higher gamma default mode network connectivity and poorer attention at baseline showed greater response to tDCS. Higher E-fields were associated with greater response. No serious adverse effects occurred.

**Conclusions:** A **s**ingle tDCS session targeting dlPFC or IFG did not improve attention following pediatric ABI. We demonstrated how HD-EEG source-based connectivity may be used to personalise tDCS. Future research should explore whether personalization, and/or repeated tDCS sessions can improve attention following pediatric ABI.

## Introduction

Acquired brain injury (ABI), defined as any injury occurring to the brain since birth, is a leading cause of death and lifelong disability in children.^1,2^ ABI can have life-long neurological, psychological, and cognitive sequelae which impair learning and development in childhood and adolescence, affecting later educational attainment, career opportunities and community participation.^3^ One of the most commonly-reported cognitive problems following ABI is attention deficit, and in particular, inhibitory control^4–6^, where up to 20% of children are diagnosed with attention deficit hyperactivity disorder (ADHD) after TBI^7^, and up to 46% following childhood stroke.^8^ As a component of attention, inhibitory control relates to the ability to engage in an appropriate attentional focus while suppressing a more dominant, but less appropriate, focus.^9^ Disruptions to prefrontal attention networks such as the executive control network (ECN) and salience network (SN) are thought to underpin attention deficits following ABI.^10,11^

Although stimulant medications are the mainstay treatments for attention problems, patient compliance is often poor due to side effects, concerns about misuse^12^, and stigmatisation.^13^ Therefore, non-pharmacological methods to improve attention following ABI are attractive alternatives. Transcranial direct current stimulation (tDCS) has been demonstrated to improve attention in adult ABI^14^ by transiently modulating cortical activity in regions with abnormal activity.^15–18^ Two main tDCS targets have been previously studied: the left dorsolateral prefrontal cortex (ldlPFC); an important hub of the ECN^19–21^ and postulated key player in executive dysfunction following ABI^22,23^, and the right inferior frontal gyrus (rIFG); a key hub in the SN^14,21,24–32^ which is thought to underpin problems with impulsivity and inhibition in children.^33–36^ However, as key developmental changes occur in these networks during adolescence, the optimal target for tDCS in a childhood ABI population is unknown.

Inter-individual variability in tDCS response is a challenge.^37^ This is especially true following ABI due to the heterogeneous nature of injury and resultant brain network changes. Brain connectivity changes have been traditionally measured using MRI^23,38,39^, but more recently with HD-EEG^40–42^, which has the advantage of being more child-friendly, cost-effective, and portable, facilitating use at the point of care.^43,44,45^ Further, injury-related differences in tDCS-induced cortical electric field can occur due to secondary gliosis, leukomalacia and/or cortical atrophy, leading to differences in response.^46–48^ This has been seen in adults with ABI where greater SN structural integrity was associated with a greater response to tDCS^49^, indicating the importance of accounting for brain connectivity when assessing tDCS effects.

In this study, we investigated the use of tDCS targeting the dlPFC or IFG during gamified training to improve attentional inhibition in children with ABI. To address the challenge of inter-individual variability in tDCS response, we also investigated the association of structural connectivity (sc), functional connectivity (fc), baseline attention, and simulated cortical E-fields with tDCS response. We hypothesised that tDCS would be feasible and tolerable, and that active tDCS (targeting dlPFC or IFG) would result in greater attention improvements compared to sham tDCS. Finally, we hypothesised that tDCS-related attention improvement would be greatest in those with greater sc and fc in attention networks, greater baseline attention, and with higher simulated E-field in prefrontal regions.

## Methods

Our study investigated whether a single session of 1 mA tDCS during attention training significantly improved attention when applied over the bilateral dlPFC or bilateral IFG in children and adolescents in the chronic stage of ABI and similar healthy controls (HCs). This pre-registered, randomised, sham-controlled trial was conducted at the University of Queensland Child Health Research Centre (Brisbane, Australia) between November 2021 and December 2022 (OSF registration: <u>osf.io/b43nh</u>). Fifteen participants with ABI between the ages of 7-18 years (inclusive) and 15 healthy controls (HCs) participated in the study. Three single sessions of tDCS were given (left dlPFC anode/right dlPFC cathode, right IFG anode/left IFG cathode, or sham), separated by >48 hours (Fig. 1). Baseline assessments included HD-EEG, MRI (optional) and clinical/neurocognitive assessments. Change in flanker reaction time (RT) or stop signal reaction time (SSRT) was used to measure changes in attention following tDCS.^50–52^ Any short-acting ADHD medication was temporarily discontinued 48 hours prior to study. The study was conducted in accordance with Good Clinical Practice and ethical approval was received by the University of Queensland (HREC/21/QCHQ/73034). All participants gave written assent and a parent/guardian gave written consent for participation in the study.

**Figure 1:**
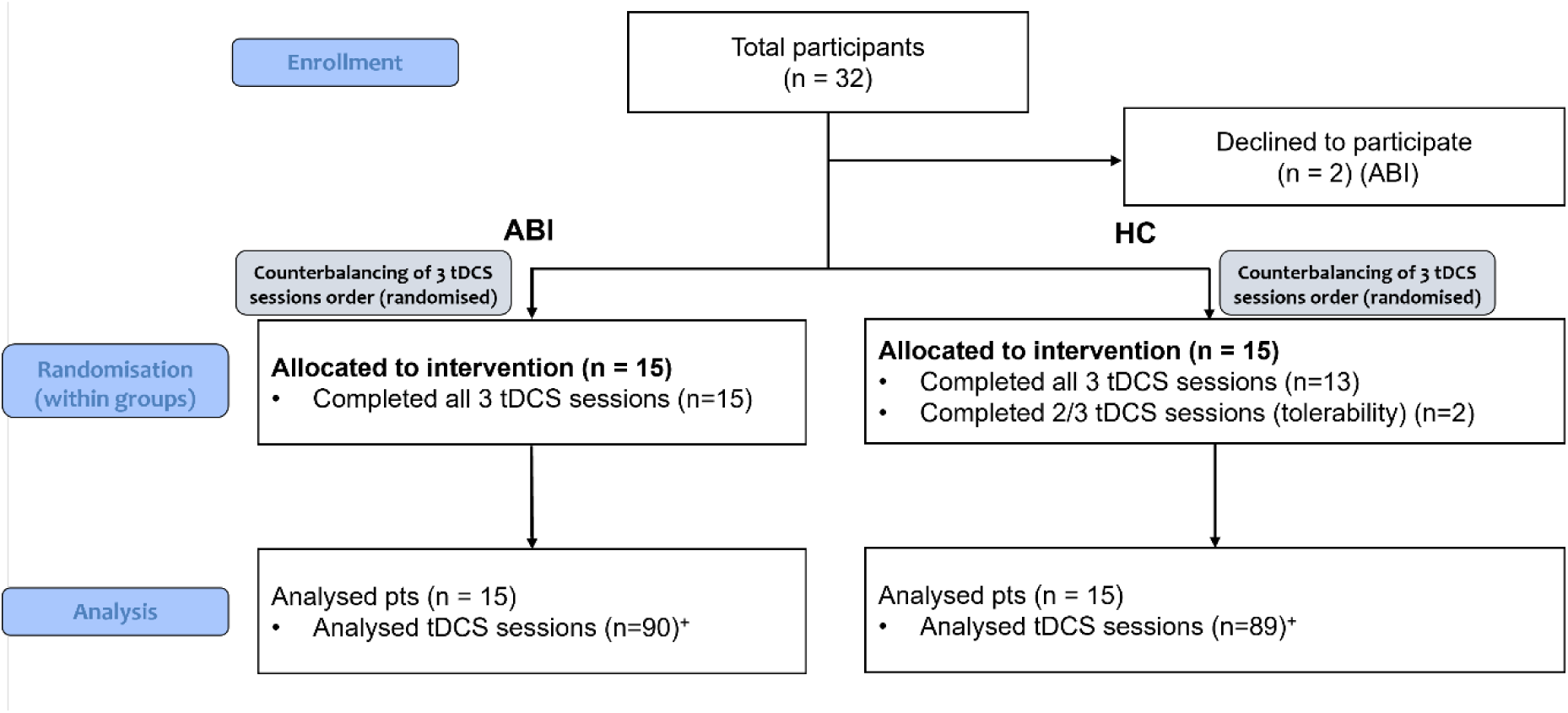
CONSORT flow diagram. tDCS, transcranial direct current stimulation * Out of a total of 90 tDCS sessions

### Inclusion and exclusion criteria

Participants were eligible if they were between 7 and 18 years old (inclusive), and ABI participants were >3 months post-injury (mild TBI) or >1 year post injury (moderate-severe TBI). ABI was defined as any injury to the brain occurring after birth.^1^ Exclusion criteria included contraindications to tDCS, hypoxic-ischemic encephalopathy, significant past medical or psychiatric history before injury, recent or planned change in neuroactive drugs, pre-existing neurological disorders including moderate to severe learning difficulties, or presence of a condition that in the opinion of the Investigator would compromise the safety of the patient or quality of the data. See Supplementary Methods for definition of ABI and more detail about these exclusion criteria.

#### Sample size, randomisation, and blinding

Based on previous work in adult ABI^14^, a sample size of 13 participants per group would allow detection of a moderate effect (Cohen’s d=0.69) using EEG changes. To compare group differences in flanker RT or SSRT, a Bayesian stopping rule was set at BF_10_>3. Maximum group sample size was set at n=15. The session order was counterbalanced using computer-generated block randomisation sequences (block size 6). Participants and families only were blinded to the intervention.

### Outcome measures

The primary outcome was change in flanker^53^ RT using the NIH toolbox (iPad).^54^ This 3 minute task consists of 24 congruent and incongruent trials. The flanker task has strong validity and test-retest reliability in paediatric TBI populations.^55^ Change in RT was also assessed using a gamified Stop Signal Task (the ‘Fairy Game’^56^) consisting of 200 trials (148 Go and 52 Stop trials) across two 6 minute blocks with a 15 second break. This game has demonstrated validity compared to the traditional SST^56^, has been used in children with ADHD^57^, and is sensitive to change with dlPFC tDCS^58^ (see Supplementary Methods for further details). Safety and tolerability were assessed immediately following each session using the Sensations/Adverse Effects Questionnaire (Supplementary Figure 1).

### Intervention

Each participant received three single sessions of tDCS (two active, one sham) separated >48 hours using a counterbalanced, within-subjects design. Using the 10/20 EEG system^18^, the anodal stimulation site was F3 (left dlPFC [ldlPFC]) or F8 (right IFG [rIFG]). The cathode was placed contralaterally on F4 (right dlPFC) or F7 (left IFG), respectively. The ldlPFC was chosen as a target for executive control of attention^19,21^ and rIFG as a target for inhibitory control.^30,36^ For the sham condition, participants were equally randomised to a sham ldlPFC or sham rIFG montage. Active stimulation was delivered for 20 minutes at 1mA using 25cm^2^ saline-soaked sponge electrodes (NeuroConn DC Stimulator Mobile, Ilmenau, Germany), with a calculated current density of 0.04 mA/cm^2^ per electrode. Ramp-up and ramp-down time was 30s. Sham stimulation involved 30 seconds of ramp-up followed by immediate 30 seconds ramp-down.^59^ During stimulation, participants completed gamified attention training (16 minutes, Go/No-Go task: ‘Wormy Fruit’, and continuous performance task: ‘SnappyApp’; see Supplementary Methods).^60^

### Baseline assessments

Cognitive function was assessed using a computerised neuropsychological assessment (CNS Vital Signs [CNSVS]), which has demonstrated validity in this population.^61^ The Behaviour Rating Inventory for Executive Functioning (BRIEF2 Parent Report, Second Edition) was used to assess executive functioning and likelihood of ADHD.

### Structural and anatomical MRI

#### MRI acquisition

Structural MRI was acquired using a 3T Siemens Prisma scanner with a 64-channel head coil at the Herston Imaging Research Facility (HIRF; Brisbane, Australia). MRI was used to assess anatomy (T1, T2), sc (DTI) and to perform electric field modelling in the ABI participants.

#### Electric field modelling

Individual head models (n=12) were created for ABI participants from T1 and T2 scans (n=9) or T1 scans only (n=3) using the headreco segmentation and meshing pipeline (Copenhagen, Denmark)^62,63^ in MATLAB (R2021a, Massachusetts: The MathWorks, Inc). Following segmentation, head models were converted to SimNIBS 4.0.0. where electric field (E-field) models were created for each montage.^64^ The ldlPFC/right dlPFC montage was modelled according to F3-anode/F4-cathode (10-20 EEG), and the rIFG/left IFG montage was modelled according to F8-anode/F7-cathode (electrode size:5×5cm; 4 mm thickness). Electrode y-axis pointed towards FC3 and FC4 for F3 and F4 electrodes, respectively. Current intensity was 1 mA/-1mA for anode and cathode, and default conductivity was utilized.

Mean E-field magnitude (E_norm, V/m) was calculated in a 10 mm spherical radius ROI with a grey matter mask centred at the cortical projection underneath the anodal and cathodal electrodes at the following MNI locations: F3 (-35.5, 49.4, 32.4), F4 (40.2, 47.6, 32.1), F7 (-54.8, 33.9, -3.5), and F8 (56.6, 30.8, -4.1).^64–66^ Sham E-field was calculated by dividing the calculated electric field for each participant by 20 due to the linear relationship between active and sham tDCS. The E-field distribution map was visualised using gmsh.^67^

#### Tractography

Details about DTI preprocessing and model fitting are given in the Supplementary Methods. Tracts of interest underlying the salience (SN) and default mode (DMN) networks were used to examine associations between sc and RT change following tDCS^49^ including the frontal aslant tract (FAT)^68–70^, and the cingulum bundle (CG)^71^ (Supplementary Figure 2). Mean FD and FA were extracted from all tracts of interest for subsequent correlation with tDCS response.

### Functional connectivity

#### HD-EEG Acquisition

High-density electroencephalography (HD-EEG) was used to assess fc at baseline using a 128 channel cap (Geodesic EGI Hydrocel GSN 130), during 5 minutes of resting eyes open (EO) while watching ‘Inscapes’^72^ and 5 minutes of resting eyes closed (EC), sampled at 250 Hz using Net Station (Version 5.4.2, Philips Electrical Geodesics, Eugene, Oregon). Impedance was kept below 100 kΩ.

#### Source-based connectivity model

The HD-EEG pre-processing pipeline is outlined in Figure 2 and Supplementary Methods. The Brainstorm toolbox (Version February 2023)^73^ was used for source-based connectivity analysis. Where available, T1 MRI scans were used for subject anatomy (n=12 ABI participants). Alternatively, NIHPD age-specific asymmetric T1 templates were used (n=15 HC participants and n=3 ABI participants).^74^ CAT12^75^ was used for T1 segmentation and EEG channel files were registered onto MRI-based head models. Boundary element model (BEM)^76^ surfaces were generated using default Brainstorm settings to define conductor models in forward model computation. Diagonal noise covariance was computed (default identity matrix) and head models were created (15002 vertices, adjoint formulation). Forward model source reconstruction was conducted using dynamical Statistical Parametric Mapping (dPSM)^77^ with single kernel.

**Figure 2:**
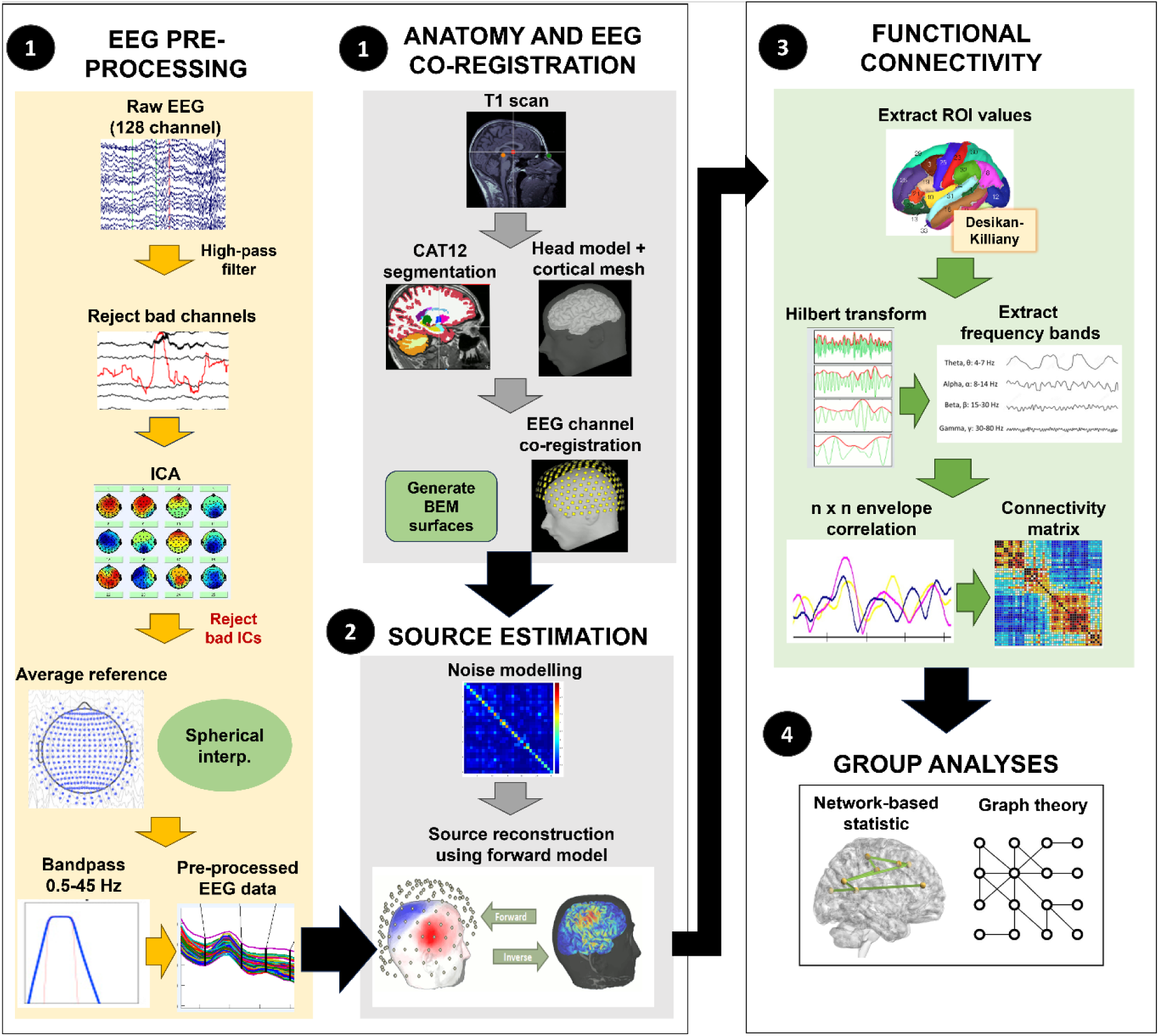
HD-EEG preprocessing and functional connectivity analysis pipeline. HD-EEG, high-density electroencephalography; ICA, independent components analysis; IC, independent components; CAT, computational anatomy toolbox; BEM, boundary element method; ROI, region of interest

#### Functional connectivity

To analyse connectivity, the cortex was parcellated according to the Desikan-Killiany Atlas^78^, and the source-reconstructed time series was computed across 68 cortical regions. Orthogonalised envelope correlation was used to measure brain connectivity across frequency bands (Hilbert time-frequency transformation; delta=1-3.99 Hz, theta=4-7.99Hz, alpha=8-12.99Hz, beta=13-30Hz, gamma=30.01-40Hz, and broadband=1-40Hz; static time resolution; sliding window length=1500ms with 50% overlap).^79^ Significant group differences in subnetwork connectivity were identified using network-based statistics (NBS).^80^ Region of interest-based analyses were also conducted within the DMN, SN and ECN (See Supplementary Methods).

### Statistical analysis

Statistical analysis and visualisation were performed using MATLAB Version 9.12 (R2022a, The MathWorks Inc, Natick, Massachusetts), RStudio (Version 2023.9.1.494, PBC, Boston, MA), Just Another Statistics Program (JASP; Version 0.16.2.0 for Windows), Graphpad Prism Version 9.0.0 (GraphPad Software, Boston, Massachusetts) and Statistical Package for the Social Sciences (SPSS; IBM, Armonk, NY). Flanker trials were excluded if RT<100 ms or the response was incorrect.^81^ SSRT was calculated using the Verbruggen method.^82^ Trials were excluded if the accuracy rate was <80%, if RT on unsuccessful stop trials was longer than RT on go trials (race model violation), or if probability of responding on a stop signal was <0.25 or >0.75.^83,84^ Non-normal RT data was not transformed as per Lo & Andrews. 2015^85^. Post-hoc analysis of outliers was conducted using the extreme studentized deviate (Grubbs) test.^86^

The primary outcome (post-tDCS flanker RT) was assessed in each group (ABI, HC) using a series of liner mixed models (LMM), controlling for pre-tDCS flanker RT and intervention arm (dlPFC, IFG, sham) as fixed effects, and participant ID as a random effect. A between-group model was also performed with a fixed effect of group. Post-hoc, session order was added as a fixed effect. A random intercept model was used with sham condition as the reference category. Details of statistical models are given in Supplementary Equations 1 and 2. As a secondary outcome, change in SSRT across interventions was compared using repeated-measures ANOVA in each group separately with time as a repeated factor and a between factor of intervention. Finally, group differences were assessed by adding group as a between-subjects factor.

To test for significant differences in baseline fc between groups, we performed group-wise contrasts including with age as a covariate. NBS tests were conducted with 5000 permutations at p=0.05. Comparisons were made across delta, theta, alpha, beta, gamma, and broadband frequencies in EC and EO conditions at a t-statistic threshold of 3.1 or 3.8 (expected medium effect size between 0.5 and 0.6). For network ROIs (ECN, SN, DMN), independent two-sample t-tests were conducted (two-tailed, p<0.05) to investigate differences in connectivity in each ROI between groups. Correlation analysis was used to examine the association between baseline attention performance and baseline fc. Similarly, we also examined associations between baseline attention performance and baseline structural connectivity (FA, FD).

#### Potential factors affecting tDCS response

In a series of exploratory analyses, we examined the effect of ABI-related anatomical variation on outcome. Correlations were used to examine the association between RT change following dlPFC or IFG tDCS and baseline fc (n=15 ABI, n=15 HC), structural connectivity (n=10 ABI), attention (n=15 ABI, n=15 HC), and magnitude of simulated anodal E-field (n=12 ABI). The association between E-field and microstructural injury (FA, FD) was also examined. As a post-hoc analysis, we assessed the effect of age on the factors listed above. Multiple comparison correction was not performed due to the exploratory nature of the analyses.^87^

## Results

### Participants

Demographic and clinical details of participants are reported in Table 1. Mild TBI was the commonest cause of injury (TBI n=13, stroke n=1, meningitis n=1). Baseline cognitive function did not significantly differ between groups. Behavioural attention was significantly poorer in the ABI group (see Table 1). Baseline flanker RT was not significantly different between groups (t=-0.83, p=0.41; see Supplementary Table 1 for values across tDCS arms). As expected, baseline flanker RT correlated with age in both the ABI (r=-0.86, p<0.001) and HC groups (r=-0.72, p=0.002), where older participants had faster RT.

**Table 1:** Baseline demographic, neurocognitive, behavioural, and quality of life measures.

| Measure | HC | ABI | Test statistic | p val |
| --- | --- | --- | --- | --- |
| Age (y), mean (SD, 95% CI) | 11.41 (2.87, 9.82-13.00) | 12.68 (3.33, 10.84-14.53) | t=-1.12 | 0.27 |
| Sex, n males (%) | 9 (60.00) | 11 (73.3) | $\chi^2=0.60$ | 0.70 |
| Handedness, n right (%) | 12 (80.00) | 14 (93.33) | $\chi^2=1.15$ | 0.27 |
| Time post-injury (y), mean (SD, range) | NA | 3.81 (2.74, range 1-9 y) | NA | NA |
| Injury severity |  |  | NA | NA |
| Mild | NA | 10 |  |  |
| Moderate | NA | 2 |  |  |
| Severe | NA | 3 |  |  |
| CNSVS NCI, mean (95% CI) | 96.73 (91.22, 102.25) | 91.00 (83.38, 98.63) | t=1.31 | 0.20 |
| CNSVS RT, mean (95% CI) | 94.80 (83.62, 105.98) | 93.47 (86.27, 100.66) | t=0.22 | 0.83 |
| CNSVS Simple Attention, mean (95% CI) | 83.00 (68.50, 97.50) | 64.33 (40.13, 88.54) | t=1.42 | 0.17 |
| CNSVS Complex Attention, mean (95% CI) | 94.27 (88.65, 99.88) | 77.73 (59.98, 95.48) | t=1.91 | 0.07 |
| BRIEF2 BRI, mean (95% CI) | 47.47 (44.19, 50.74) | 64.93 (57.52, 72.34) | t=-4.62 | **<0.001** |
| BRIEF2 Inhibit, mean (95% CI) | 48.13 (44.20, 52.07) | 62.93 (55.53, 70.34) | t=-3.79 | **0.001** |
| BRIEF2 Shift, mean (95% CI) | 47.80 (43.63, 51.97) | 73.00 (64.92, 81.08) | t=-5.95 | **<0.001** |

SD, standard deviation; CNSVS, CNS Vital Signs; NCI, Neurocognitive Index; RT, reaction time; BRI, Behaviour Regulation Index; PedsQL, Pediatric Quality of Life Scale; CHU9D, Child Health Utility Index 9-Dimensions; CI, confidence interval

### RT change following intervention

One session (HC) was incomplete (less than half of session completed) and excluded. A total of 89 sessions (out of 90) were included in the analysis. There was no effect of intervention on post-intervention flanker RT in either group (ABI: F=0.40, p=0.67; HC: F=0.55, p=0.58; Fig. 3, Supplementary Tables 2-4). Less RT change was seen in ABI participants with faster baseline RT (F=25.11, df=1064.3, p<0.001; Supplementary Table 3). Post-intervention flanker RT did not significantly differ between groups (ABI vs. HC) across interventions, after controlling for baseline flanker RT (F=0.66, p=0.42). There was no significant effect of tDCS condition on SSRT (Supplementary Table 5, Supplementary Figure 3). Slower baseline flanker RT was correlated with greater flanker RT change (i.e., better RT) in the dlPFC tDCS condition only. Baseline flanker RT was not associated with flanker RT change in the IFG or sham tDCS conditions (Supplementary Figure 5).

**Figure 3:**
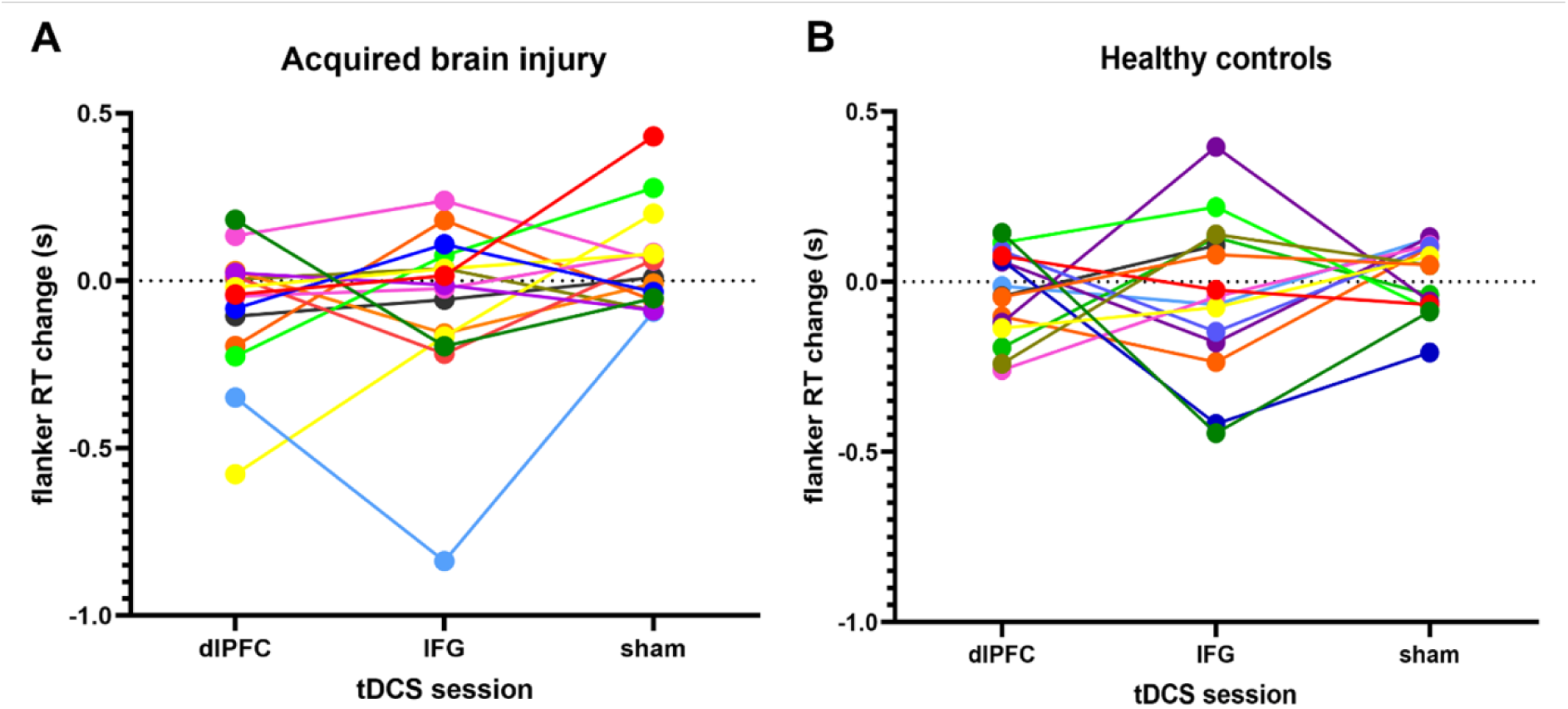
Mean flanker RT change across tDCS arm (dlPFC, IFG, sham) in ABI and HC groups. Colours represent within-subject data. RT, reaction time; tDCS, transcranial direct current stimulation; s, seconds; dlPFC, dorsolateral prefrontal cortex; IFG, inferior frontal gyrus; ABI, acquired brain injury; HC, healthy control

### Baseline connectivity and attention

Significant between-group differences were identified in several subnetworks across spectral bands (delta, theta, alpha, beta, gamma, broadband) and are demonstrated in Figure 4. In the EO condition, ABI participants showed significantly increased delta connectivity in an insula-centric subnetwork connecting to parietal DMN nodes and another subnetwork involving the superior frontal, inferior frontal, transverse temporal, posterior cingulate and fusiform regions (t=3.1, Network 1: p_FWE_=0.013, Network 2: p_FWE_=0.012; Fig. 4A,B). In ABI participants only, this was associated with higher (i.e., slower) baseline flanker RT (Spearman’s rho =0.60, p=0.02; Fig. 4C). ABI participants also had increased theta connectivity in the EO condition in a fronto-parietal subnetwork compared to HCs (t=3.1, p_FWE_=0.016; Fig. 4D,E). In the EC condition, ABI participants had significantly decreased alpha connectivity in a subnetwork of nodes across fronto-parietal DMN regions (age corrected, t=3.8, p_FWE_=0.003; Fig. 4F,G), and significantly increased gamma in another fronto-parietal DMN-like subnetwork (age corrected, t=3.1, p_FWE_=0.009; Fig. 4H,I) compared to HCs. Significant nodes in each network are listed in Supplementary Table 6. There were no significant group differences in ROI connectivity (ECN, DMN or SN) or power spectra in delta, theta, alpha, beta, gamma, or broad bands, and no associations between connectivity and baseline SSRT. Greater structural connectivity (increased FA of the right FAT) was associated with faster flanker RT in ABI participants (see Supplementary Figure 4).

**Figure 4:**
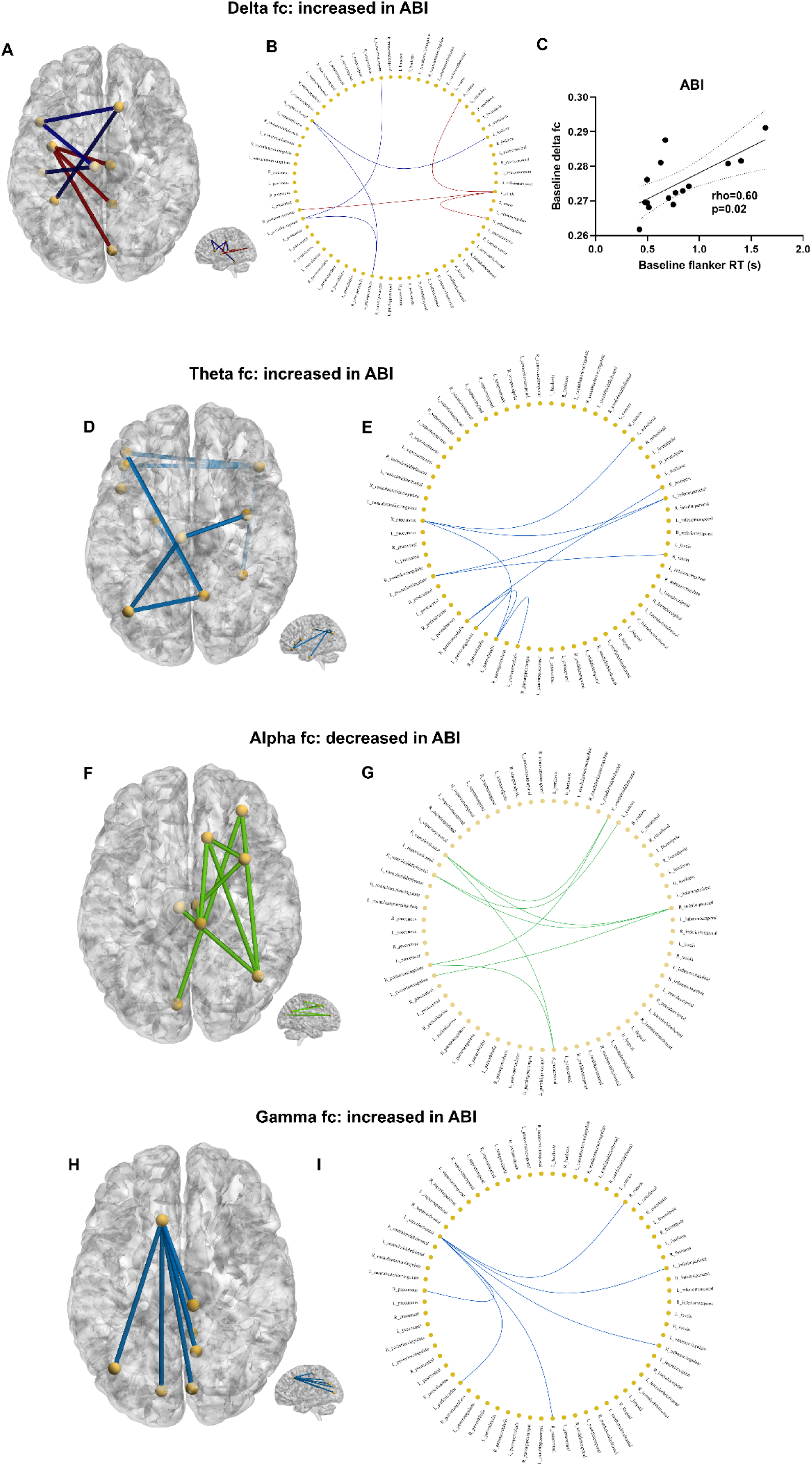
NBS subnetworks showing significant differences in connectivity in ABI participants compared to HCs at baseline and associations with baseline flanker RT. Increased resting EO delta connectivity in ABI participants displayed as (A) nodes and edges on brain template or (B) in a circular graph. (C) Increased delta connectivity associated with slower flanker RT at baseline in ABI participants. Increased resting EO theta connectivity in ABI participants displayed as (D) nodes and edges on brain template or (E) in a circular graph. Decreased resting EC alpha connectivity in ABI participants displayed as (F) nodes and edges on brain template or (G) in a circular graph. Increased resting EC gamma connectivity in ABI participants displayed as (H) nodes and edges on brain template or (I) in a circular graph. No significant associations between connectivity and attention in theta, alpha, beta, gamma or broad band subnetworks. NBS t-tests used to determine significant subnetworks. All networks corrected for age. t=3.1 in delta, theta and gamma, and t=3.8 in alpha network. Edge colours represent separate significant subnetworks. Displayed using BrainNet Viewer and NeuroMarvl. NBS, network-based statistics; s, seconds; RT, reaction time; ABI, acquired brain injury; HC, healthy controls; EO, eyes open; EC, eyes closed.

### Factors affecting tDCS response in children with ABI

#### Functional connectivity

Both with and without a consistent outlier that was finally removed from analysis, greater (faster) flanker RT change following IFG tDCS was associated with lower theta (slow wave) fc in DMN (ROI analysis), and greater (faster) SSRT change following dlPFC tDCS was associated with higher gamma fc in DMN (Figure 5). There were no other significant associations and ROI fc was not associated with age.

**Figure 5:**
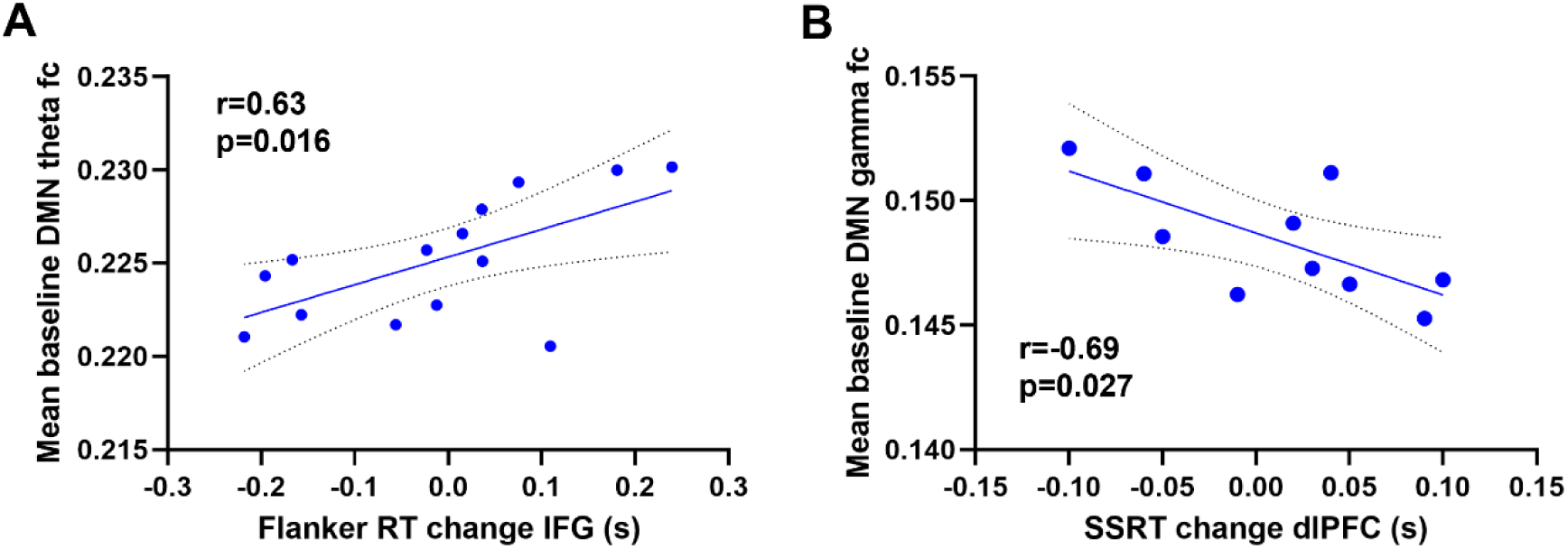
Significant correlations between mean baseline functional connectivity using HD-EEG (resting EC) and mean RT change following tDCS in ABI participants after excluding persistent outlying participant. (A) Lower baseline theta DMN connectivity was associated with faster flanker RT following IFG tDCS. (B) Higher baseline gamma DMN connectivity was associated with faster SSRT following dlPFC tDCS. Pearsons’s correlations, 95% confidence intervals displayed. tDCS, transcranial direct current stimulation; RT, reaction time; SSRT, Stop Signal Reaction Time; DMN, default mode network; s, seconds; dlPFC, dorsolateral prefrontal cortex; IFG, inferior frontal gyrus; ABI, acquired brain injury; EC, eyes closed.

#### E-Field and structural connectivity

Mean normalised E-field in the anodal target regions were 0.211 (SD 0.05) V/m in the ldlPFC, 0.209 (SD 0.07) V/m in the rIFG and 0.011 (SD 0.01) V/m in the sham condition (Figure 6). E-field was not correlated with age in either the dlPFC (r=-0.56, p=0.06) or IFG (r=0.13, p=0.69) conditions.

**Figure 6:**
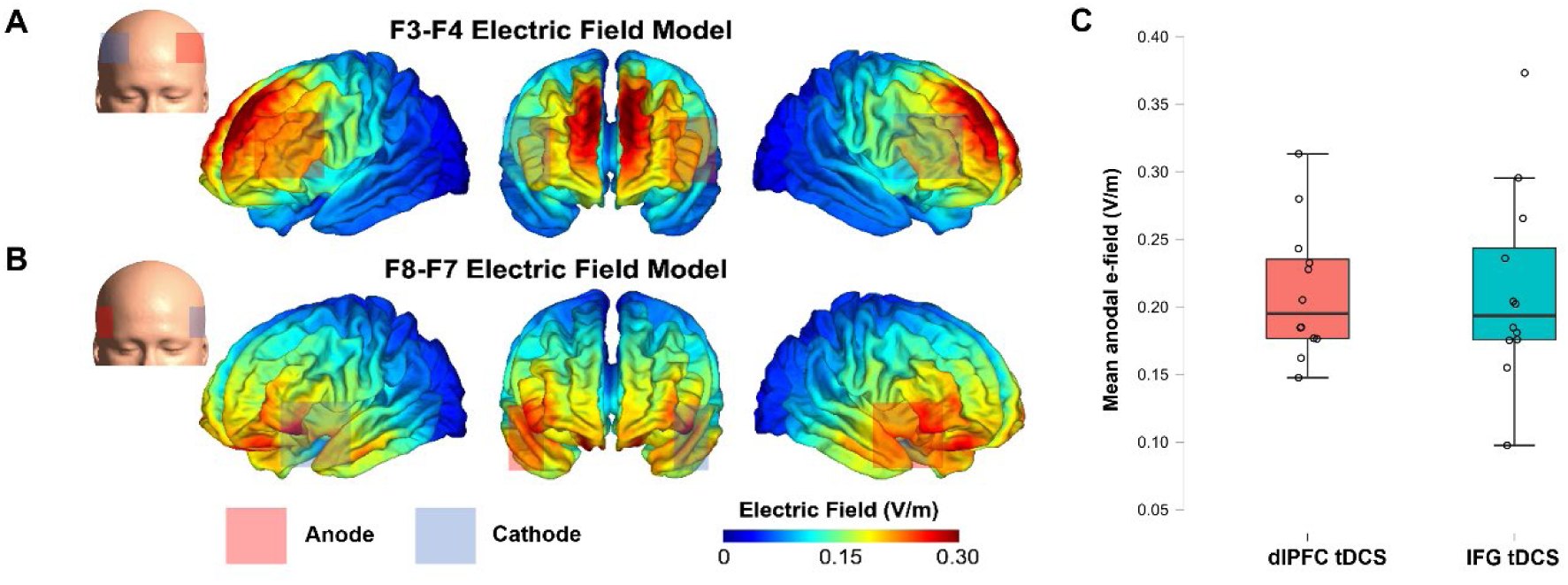
Mean normalised electric field across ABI participants in active tDCS sessions. (A) Visual representation of tDCS montage in dlPFC (F3/F4) and (B) IFG arms (F8/F7). (C) Mean normalised electric field across participants in dlPFC and IFG tDCS (n=12). dlPFC, dorsolateral prefrontal cortex; IFG, inferior frontal gyrus; tDCS; transcranial direct current stimulation; E-field, electric field; RT, reaction time; s, seconds

Participants with higher E-field at the ldlPFC showed faster flanker RT following the ldlPFC tDCS intervention (rho=-0.60, p=0.04; Supplementary Figure 6). No other significant associations were found between E-field and flanker RT change following the IFG tDCS or sham conditions, or between E-field and SSRT change.

Baseline sc (FA or FD) was not associated with flanker RT or SSRT change in any ROI across interventions. Right-hemisphere FA and FD in both the FAT and CG was positively associated with age. E-fields were not associated with structural connectivity.

### Tolerability and blinding

Most participant sessions (n=88) were tolerated well, and only minor adverse effects were reported. No differences in tolerability were seen between groups (Fig. 7). Two HC participants (8 and 9 years old) were unable to tolerate one session each (IFG and sham) due to itching and discomfort. Both participants, however, successfully completed the remaining tDCS sessions. The most common sensation was mild itching and did not differ between groups (χ^2^=1.4, p=0.75). Reports of mild pain during tDCS did not differ between groups (ABI, 4%; HC, 7%; t=2.04, p=0.49). One ABI participant retrospectively reported the pain as strong, yet they were not distressed during the session and wished to complete the session (Fig. 7C). Blinding was successful as the majority of participants incorrectly guessed that they received sham, and there were no significant differences in correct sham guess between groups (χ^2^= 0.59, p=0.70) (Figure 7D).

**Figure 7:**
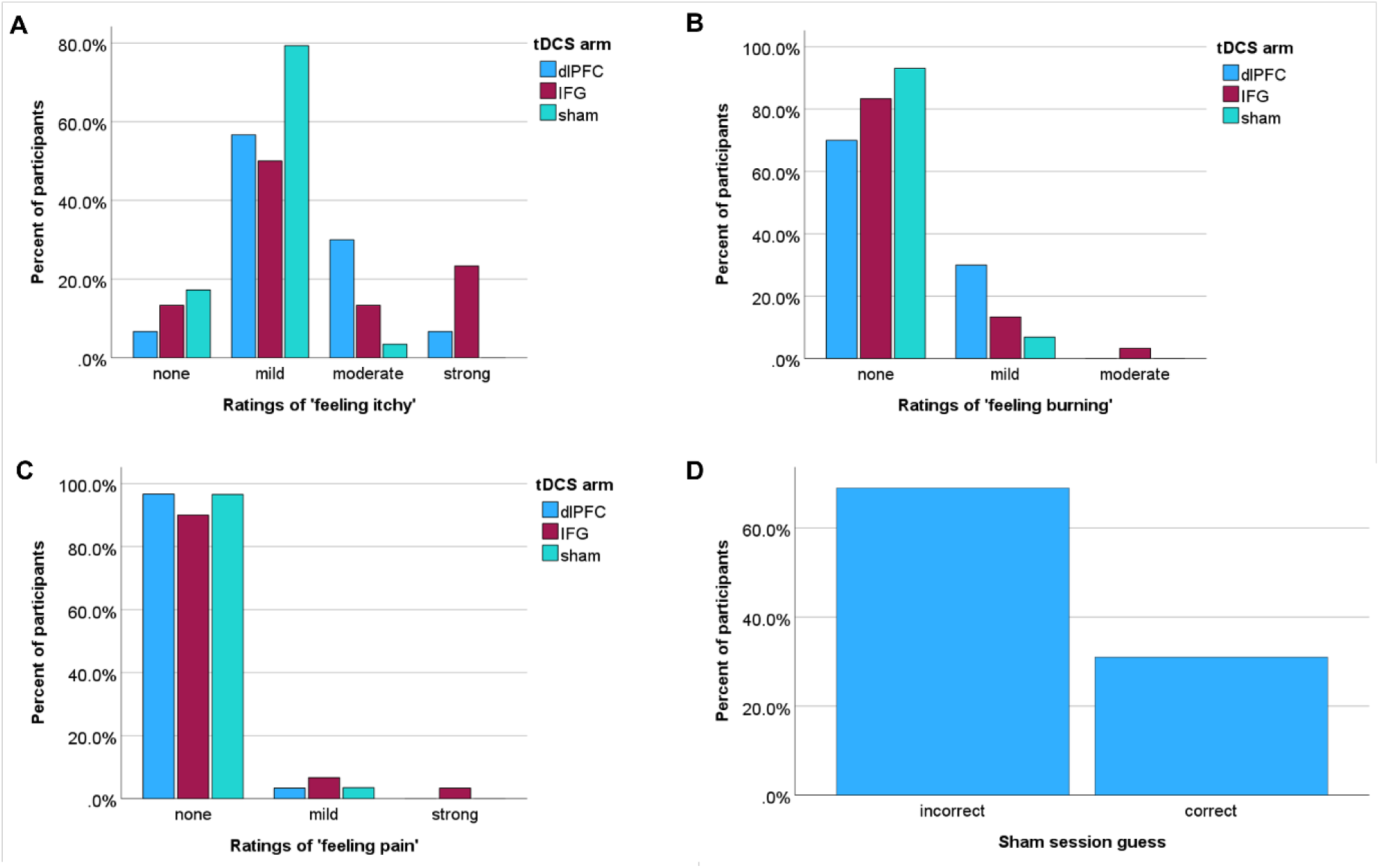
Percentage of patients reporting severities of (A) ‘itchy’, (B) ‘burning’ and (C) ‘pain’ sensations (ABI and HC) following each tDCS session. Sensation severity ratings did not differ across tDCS arms or between study groups. (D) Percentage of participants who correctly or incorrectly guessed that they received sham stimulation in the sham tDCS session tDCS, transcranial direct current stimulation; dlPFC, dorsolateral prefrontal cortex; IFG, inferior frontal gyrus

## Discussion

Attention problems following childhood ABI cause life-long disruptions in functioning and quality of life.^1–3^ Pharmacological therapies are effective, but are associated with significant side effects.^12,13^ In this study, we investigated the efficacy and feasibility of tDCS during attention training as a potential alternative to treat attention problems in children with acquired brain injury. We evaluated two common stimulation sites for attention (dlPFC and IFG, as hubs for the ECN and SN, respectively) given the potential for differences in attention network organisation across childhood development to influence tDCS response.^88^ We also addressed the challenge of inter-individual variability in response in this heterogenous population through a multi-modal analysis investigating whether fc, structural connectivity, baseline attention, or simulated E-fields were associated with response to the tDCS intervention. Overall, we found that 1 mA was tolerable in children with ABI. A single session of tDCS during gamified attention training did not significantly improve attention measured using flanker or stop signal reaction times when compared to sham tDCS, in either ABI or HC participants. However, we found several sources of inter-individual variability which may have affected tDCS response.

Although children with ABI in our study had attention-related disruptions in functional brain connectivity, there was no effect of a single tDCS session on attention when delivered to the dlPFC or IFG. The lack of a robust effect of a single tDCS session has been reported previously, albeit in other populations.^26,89–99^ Our small sample size may well have influenced our ability to demonstrate any effect.^100^ More recently, the utility of multiple tDCS sessions to facilitate the neuroplastic changes required for a robust response to tDCS has been reported in healthy populations, adults with ABI, and in children and adults with ADHD.^14,25,101–105^ When applied repeatedly (i.e., over the course of a day or several days), the effects of tDCS are cumulative.^104,105^ Our null results could also have reflected suboptimal brain targeting in a childhood population as attention networks shift from a rostral to caudal distribution, mirroring the shift from dependence on lower-level sensory areas to complex association areas.^88,106–108^ It is also important to consider that variability in tDCS response may have occurred due to inter-individual variability arising from injury, ongoing development or other comorbid diagnoses.^37^ In the future, measurement of EEG following the intervention should be included to provide greater insight into neurophysiological changes.

A unique element of our study was the use of HD-EEG to measure source-derived fc at baseline, which provided a simple, child-friendly measure of brain connectivity.^44,45,109^ We found that children with ABI had poorer attention before the intervention which correlated strongly with evidence of frontoparietal network dysfunction (increased slow wave connectivity). This, at least in part, supports our use of prefrontal tDCS to modulate attention.^41,110–114^ We also found evidence of DMN hyperactivity in children with ABI before the intervention, including decreases in fronto-parietal alpha connectivity and increases in gamma connectivity in a DMN-like network. This result is similar to previous fMRI research which has found DMN hyperactivity following pediatric ABI to be a hallmark of injury-related executive dysfunction.^23,42,115–123^ Our hypothesis that increased functional network connectivity following ABI would relate to greater tDCS response was supported, as participants with greater gamma (fast wave) connectivity and lower theta (slow wave) connectivity in the DMN showed greater improvements in reaction time following tDCS. While understanding of frequency-dependent connectivity changes following ABI is limited, this suggests that DMN connectivity may explain some of the inter-individual variability in tDCS response. However, as this association was derived from fc measured from pre-defined ROIs in patients only, it is not clear whether this relationship is purely due to injury factors, as we did not find differences in ROI connectivity in this network between ABI and HC participants. Therefore, it seems likely that abnormal DMN activity may influence response to tDCS in children with ABI^11,23,123,124^; however, future research is needed to confirm this finding.

Interestingly, those ABI participants with poorer attention at baseline had the greatest improvement in attention following dlPFC tDCS. This is in keeping with previous literature and may potentially explain the lack of overt response to tDCS in our sample as most had reaction times within the normal range. Participants with faster RT prior to the intervention demonstrated less change following the intervention, perhaps because they had less ‘room to improve’ compared to those with a slower RT.^125–128^ This phenomenon has been reported in a tDCS trial in adults with TBI^14^, as well as in healthy adults in both the cognitive^129–131^ and motor domains.^128^ We suggest that future research should consider baseline attention as a potential mediator of tDCS response, as participants with attention deficit may benefit most greatly from tDCS intervention.^23^

Heterogeneity in severity and extent of brain tissue injury affects tDCS current flow and magnitude, leading to variability in tDCS response.^46,132,133^ We hypothesised that our participants would have variable cortical E-fields, despite receiving the same ‘scalp intensity’ of tDCS, and that this could contribute to inter-individual differences in tDCS response. Supporting our hypothesis, we found a moderate E-field ‘cortical dose’-response relationship where higher ldlPFC E-field was associated with greater RT improvement following the dlPFC tDCS condition. This relationship was not seen in the IFG condition despite similar mean E-fields. Although inhibitory control has been modulated using SN-targeted tDCS (i.e., IFG tDCS) in healthy populations^134,135^, in an ABI population, tDCS targeting the ECN (i.e., dlPFC tDCS) may be more effective in improving attention as it may address a more ‘top-down’ control of attention, in line with the executive dysfunction hypothesis in ABI.^23^ We suggest that this dose-response relationship may go some way to explain the null results of tDCS across our study population, where participants on the lower end of the dose-response (i.e., those with the lowest E-fields) may not have received a high enough cortical dose of tDCS to result in cognitive changes.^136,137^ This should be considered in the context that higher intensities of tDCS do not necessarily produce linearly greater effects, suggesting that a blanket approach of increased dose is not correct, and instead may require some personalisation.^138^ Further research using large sample sizes is needed to determine the optimal tDCS dose for attention improvement in populations with underlying anatomical/fc differences.

We also explored whether the relationship between structural connectivity, E-field variability and response to tDCS. E-field variability in brain injury populations may be caused by differences in tissue conductivity following the injury and resultant changes in structural integrity.^46,132,133,139^ In our small sample, consisting of children with relatively mild injuries (and therefore less structural damage^140^), we failed to demonstrate an association between structural integrity and E-field or RT change in children with ABI, unlike a previous tDCS study in adults with moderate to severe ABI.^49^ Indeed, our E-field modelling methods may have been insensitive to the subtlety of microstructural changes seen in mild TBI.^140,141^ In the future, E-field analyses incorporating DTI data could increase sensitivity of computational models.

In keeping with the previous literature using tDCS in children^142–145^, our results indicated that most children with ABI were able to tolerate tDCS delivered at 1 mA, and that tDCS was feasible in this population. The most commonly-reported side effect in our population was tingling.^142^ Although some children verbally reported uncomfortable itchy sensations during the tDCS session, particularly near the beginning of stimulation, they were not distressed, wished to continue the session, and most retrospectively reported ‘mild’ sensations following completion of sessions. If higher stimulation intensities in children are warranted, future research should consider employing a short tolerability test^146^ and practices to habituate participants to tDCS sensations.

Our study has several limitations. The small sample size decreases the power of the study and robustness of any conclusions. Further, our stimulation montage was bilateral, meaning that both sides of the prefrontal cortex were stimulated. This montage was used to maximise current flow, because higher peak cortical E-fields are found when electrode distance is closer (i.e., in bilateral stimulation), compared to larger distances (i.e. extra-cephalic electrodes)^66^, but may have decreased specificity of effects. The timing of tDCS sessions varied due to school commitments, which may have affected fatigue levels, reaction times and apparent tDCS response in after-school sessions.^147^ We were unable to determine the flanker congruency effect due to the limited number of flanker trials presented (NIH Toolbox)^54,148^, and future studies should ensure a greater number of flanker trials. Given the findings of this study, future research should also evaluate whether higher dosages, personalised E-field dosing, or repeated tDCS sessions have a greater effect on attention following ABI using larger sample sizes.

## Conclusions

Children with ABI tolerated 1 mA tDCS during attention training tasks well, but we failed to find any robust cognitive effect of single session of tDCS over the dlPFC or IFG on attention in children with or without brain injury. Greater tDCS response correlated with several baseline measurements, including lower theta and higher gamma-band fc in the default mode network, lower baseline attention performance, and higher strength of cortical E-fields. We demonstrated how HD-EEG source-based connectivity may be used to personalise tDCS. Further research using larger sample sizes and tDCS personalisation, particularly HD-EEG-derived connectivity, may allow for more robust characterization of potential responders to tDCS in children with ABI.

## Data Availability

The data that support the findings of this study are available on request from the corresponding author, AS. The data are not publicly available due to the sensitive nature of clinical data.

## Acknowledgements

We thank Professor Paul E. Dux for providing substantial guidance in project conceptualisation, Dr. Trish Gilholm for statistical guidance, Mr Conor Robinson for guidance with Brainstorm and NBS toolboxes, and Ms Nhi (‘Julie’) Nguyen for aid in data collection. We gratefully acknowledge the participants and their families who were involved in this study.

## CRediT Authorship statement

**Athena Stein:** Conceptualisation, Methodology, Software, Formal Analysis, Investigation, Resources, Data Curation, Writing – Original Draft, Visualisation, Project administration, Funding acquisition. **Kevin Caulfield:** Methodology, Formal Analysis, Writing – Original Draft, Visualisation. **Mervyn Singh**: Methodology, Formal analysis, Writing – Original Draft, Visualisation. **Justin Riddle:** Formal Analysis, Software, Writing – Review & Editing. **Maximilian Friehs:** Software, Methodology, Writing – Review & Editing. **Michael Craven:** Software, Methodology, Writing – Review & Editing. **Madeleine Groom:** Software, Writing – Review & Editing. **Kartik Iyer:** Conceptualisation, Methodology, Software, Formal Analysis, Writing – Review & Editing, Funding acquisition. **Karen Barlow:** Conceptualisation, Methodology, Resources, Writing – Review & Editing, Supervision, Funding acquisition.

## References

1. Greenwald BD, Burnett DM, Miller MA. Congenital and acquired brain injury. 1. Brain injury: epidemiology and pathophysiology. Archives of Physical Medicine and Rehabilitation 2003;84(3 Suppl 1):S3–7.

2. World Health Organization. Neurological disorders: public health challenges. World Health Organization; 2006.

3. Keenan HT, Bratton SL. Epidemiology and outcomes of pediatric traumatic brain injury. Developmental neuroscience 2006;28(4-5):256–63.

4. Ahmed S, Venigalla H, Mekala HM, Dar S, Hassan M, Ayub S. Traumatic brain injury and neuropsychiatric complications. Indian Journal of Psychological Medicine 2017;39(2):114.

5. Yeates KO, Armstrong K, Janusz J, Taylor HG, Wade S, Stancin T, et al. Long-term attention problems in children with traumatic brain injury. 2005;44(6):574–84.

6. Sinopoli KJ, Dennis M. Inhibitory control after traumatic brain injury in children. International Journal of Developmental Neuroscience 2012;30(3):207–15.

7. Max JE, Schachar RJ, Levin HS, Ewing-Cobbs L, Chapman SB, Dennis M, et al. Predictors of secondary attention-deficit/hyperactivity disorder in children and adolescents 6 to 24 months after traumatic brain injury. Journal of the American Academy of Child and Adolescent Psychiatry 2005;44(10):1041–9.

8. Max JE, Mathews K, Manes FF, Robertson BA, Fox PT, Lancaster JL, et al. Attention deficit hyperactivity disorder and neurocognitive correlates after childhood stroke. Journal of the International Neuropsychological Society 2003;9(6):815–29.

9. Carlson SM. Developmentally sensitive measures of executive function in preschool children. Developmental neuropsychology 2005;28(2):595–616.

10. Stuss DT. Traumatic brain injury: relation to executive dysfunction and the frontal lobes. Current opinion in neurology 2011;24(6):584–9.

11. Stein A, Thorstensen JR, Ho JM, Ashley DP, Iyer KK, Barlow KM. Attention Please! Unravelling the link between brain network connectivity and cognitive attention following acquired brain injury: A systematic review of structural and functional measures. Brain Connectivity 2023(ja).

12. Clemow DB, Walker DJ. The potential for misuse and abuse of medications in ADHD: a review. Postgraduate medicine 2014;126(5):64–81.

13. Charach A, Skyba A, Cook L, Antle BJ. Using stimulant medication for children with ADHD: what do parents say? A brief report. Journal of the Canadian Academy of Child and Adolescent Psychiatry 2006;15(2):75.

14. Ulam F, Shelton C, Richards L, Davis L, Hunter B, Fregni F, et al. Cumulative effects of transcranial direct current stimulation on EEG oscillations and attention/working memory during subacute neurorehabilitation of traumatic brain injury. Clinical Neurophysiology 2015;126(3):486–96.

15. Filmer HL, Dux PE, Mattingley JB. Applications of transcranial direct current stimulation for understanding brain function. Trends in Neurosciences 2014;37(12):742–53.

16. Nitsche MA, Paulus W. Excitability changes induced in the human motor cortex by weak transcranial direct current stimulation. The Journal of physiology 2000;527(Pt 3):633.

17. Nitsche MA, Doemkes S, Karakose T, Antal A, Liebetanz D, Lang N, et al. Shaping the effects of transcranial direct current stimulation of the human motor cortex. Journal of neurophysiology 2007;97(4):3109–17.

18. Nitsche MA, Cohen LG, Wassermann EM, Priori A, Lang N, Antal A, et al. Transcranial direct current stimulation: state of the art 2008. Brain stimulation 2008;1(3):206–23.

19. Seeley WW, Menon V, Schatzberg AF, Keller J, Glover GH, Kenna H, et al. Dissociable intrinsic connectivity networks for salience processing and executive control. Journal of Neuroscience 2007;27(9):2349–56.

20. Mesulam M-M. From sensation to cognition. Brain: a journal of neurology 1998;121(6):1013–52.

21. Menon V, D’Esposito M. The role of PFC networks in cognitive control and executive function. Neuropsychopharmacology 2022;47(1):90–103.

22. McDonald BC, Flashman LA, Saykin AJ. Executive dysfunction following traumatic brain injury: neural substrates and treatment strategies. NeuroRehabilitation 2002;17(4):333–44.

23. Sharp DJ, Scott G, Leech R. Network dysfunction after traumatic brain injury. Nature Reviews Neurology 2014;10(3):156.

24. Li LM, Violante IR, Zimmerman K, Leech R, Hampshire A, Patel M, et al. Traumatic axonal injury influences the cognitive effect of non-invasive brain stimulation. Brain 2019;142(10):3280–93.

25. Bandeira ID, Guimarães RSQ, Jagersbacher JG, Barretto TL, de Jesus-Silva JR, Santos SN, et al. Transcranial direct current stimulation in children and adolescents with attention-deficit/hyperactivity disorder (ADHD) a pilot study. Journal of Child Neurology 2016;31(7):918–24.

26. Breitling C, Zaehle T, Dannhauer M, Bonath B, Tegelbeckers J, Flechtner H-H, et al. Improving interference control in ADHD patients with transcranial direct current stimulation (tDCS). Frontiers in Cellular Neuroscience 2016;10:72.

27. Breitling-Ziegler C, Zaehle T, Wellnhofer C, Dannhauer M, Tegelbeckers J, Baumann V, et al. Effects of a five-day HD-tDCS application to the right IFG depend on current intensity: A study in children and adolescents with ADHD. Progress in Brain Research 2021;264:117–50.

28. Soff C, Sotnikova A, Christiansen H, Becker K, Siniatchkin M. Transcranial direct current stimulation improves clinical symptoms in adolescents with attention deficit hyperactivity disorder. Journal of Neural Transmission 2017;124(1):133–44.

29. Sotnikova A, Soff C, Tagliazucchi E, Becker K, Siniatchkin M. Transcranial direct current stimulation modulates neuronal networks in attention deficit hyperactivity disorder. Brain Topography 2017;30(5):656–72.

30. Aron AR. From reactive to proactive and selective control: developing a richer model for stopping inappropriate responses. Biological Psychiatry 2011;69(12):e55–e68.

31. Erika-Florence M, Leech R, Hampshire A. A functional network perspective on response inhibition and attentional control. Nature communications 2014;5(1):4073.

32. Hampshire A, Chamberlain SR, Monti MM, Duncan J, Owen AM. The role of the right inferior frontal gyrus: inhibition and attentional control. Neuroimage 2010;50(3):1313–9.

33. Bos DJ, Oranje B, Achterberg M, Vlaskamp C, Ambrosino S, de Reus MA, et al. Structural and functional connectivity in children and adolescents with and without attention deficit/hyperactivity disorder. Journal of Child Psychology and Psychiatry 2017;58(7):810–8.

34. Tremblay LK, Hammill C, Ameis SH, Bhaijiwala M, Mabbott DJ, Anagnostou E, et al. Tracking inhibitory control in youth with ADHD: a multi-modal neuroimaging approach. Frontiers in Psychiatry 2020;11:00831.

35. Konrad K, Eickhoff SB. Is the ADHD brain wired differently? A review on structural and functional connectivity in attention deficit hyperactivity disorder. Human brain mapping 2010;31(6):904–16.

36. Konrad K, Gauggel S, Manz A, Schöll M. Inhibitory control in children with traumatic brain injury (TBI) and children with attention deficit/hyperactivity disorder (ADHD). Brain Injury 2000;14(10):859–75.

37. Brunoni AR, Nitsche MA, Bolognini N, Bikson M, Wagner T, Merabet L, et al. Clinical research with transcranial direct current stimulation (tDCS): challenges and future directions. Brain stimulation 2012;5(3):175–95.

38. Jilka SR, Scott G, Ham T, Pickering A, Bonnelle V, Braga RM, et al. Damage to the salience network and interactions with the default mode network. Journal of Neuroscience 2014;34(33):10798–807.

39. Sharp DJ, Beckmann CF, Greenwood R, Kinnunen KM, Bonnelle V, De Boissezon X, et al. Default mode network functional and structural connectivity after traumatic brain injury. Brain 2011;134(8):2233–47.

40. Safar K, Zhang J, Emami Z, Gharehgazlou A, Ibrahim G, Dunkley BT. Mild traumatic brain injury is associated with dysregulated neural network functioning in children and adolescents. Brain Communications 2021;3(2):fcab044.

41. Dunkley B, Da Costa L, Bethune A, Jetly R, Pang E, Taylor M, et al. Low-frequency connectivity is associated with mild traumatic brain injury. NeuroImage: Clinical 2015;7:611–21.

42. Dunkley BT, Urban K, Da Costa L, Wong SM, Pang EW, Taylor MJ. Default mode network oscillatory coupling is increased following concussion. Frontiers in Neurology 2018;9:280.

43. Chu CJ. High density EEG—What do we have to lose? Journal of Neurophysiology 2015;126(3):433.

44. Marquetand J, Vannoni S, Carboni M, Li Hegner Y, Stier C, Braun C, et al. Reliability of magnetoencephalography and high-density electroencephalography resting-state functional connectivity metrics. Brain connectivity 2019;9(7):539–53.

45. Rolle CE, Narayan M, Wu W, Toll R, Johnson N, Caudle T, et al. Functional connectivity using high density EEG shows competitive reliability and agreement across test/retest sessions. Journal of Neuroscience Methods 2022;367:109424.

46. Stein A, Iyer KK, Barlow KM. The effect of pediatric traumatic brain injury on simulated tDCS e-field. In: New York Neuroergonomics and Neuromodulation Meeting 2022; New York, NY; 2022.

47. Datta A, Baker JM, Bikson M, Fridriksson J. Individualized model predicts brain current flow during transcranial direct-current stimulation treatment in responsive stroke patient. Brain stimulation 2011;4(3):169–74.

48. Evans C, Johnstone A, Zich C, Lee JS, Ward NS, Bestmann S. The impact of brain lesions on tDCS-induced electric fields. Scientific Reports 2023;13(1):19430.

49. Li LM, Violante IR, Zimmerman K, Leech R, Hampshire A, Patel M, et al. Traumatic axonal injury influences the cognitive effect of non-invasive brain stimulation. Brain 2019;142(10):3280–93.

50. Alderson RM, Rapport MD, Kofler MJ. Attention-deficit/hyperactivity disorder and behavioral inhibition: a meta-analytic review of the stop-signal paradigm. Journal of abnormal child psychology 2007;35:745–58.

51. Willison J, Tombaugh TN. Detecting simulation of attention deficits using reaction time tests. Archives of Clinical Neuropsychology 2006;21(1):41–52.

52. Weigard A, Heathcote A, Matzke D, Huang-Pollock C. Cognitive modeling suggests that attentional failures drive longer stop-signal reaction time estimates in attention deficit/hyperactivity disorder. Clinical Psychological Science 2019;7(4):856–72.

53. Eriksen BA, Eriksen CW. Effects of noise letters upon the identification of a target letter in a nonsearch task. Perception & Psychophysics 1974;16(1):143–9.

54. Gershon RC, Wagster MV, Hendrie HC, Fox NA, Cook KF, Nowinski CJ. NIH toolbox for assessment of neurological and behavioral function. Neurology 2013;80(11 Supplement 3):S2–S6.

55. Zelazo PD, Bauer PJ. National Institutes of Health Toolbox cognition battery (NIH Toolbox CB): Validation for children between 3 and 15 years. Wiley Hoboken, NJ; 2013.

56. Friehs MA, Dechant M, Vedress S, Frings C, Mandryk RL. Effective gamification of the stop-signal task: two controlled laboratory experiments. JMIR Serious Games 2020;8(3):e17810.

57. Gallagher R, Kessler K, Bramham J, Dechant M, Friehs MA. A proof-of-concept study exploring the effects of impulsivity on a gamified version of the stop-signal task in children. Frontiers in psychology 2023;14:1068229.

58. Friehs MA, Dechant M, Vedress S, Frings C, Mandryk RL. Shocking advantage! Improving digital game performance using non-invasive brain stimulation. International Journal of Human-Computer Studies 2021;148:102582.

59. Bikson M, Brunoni AR, Charvet LE, Clark VP, Cohen LG, Deng Z-D, et al. Rigor and reproducibility in research with transcranial electrical stimulation: an NIMH-sponsored workshop. Brain stimulation 2018;11(3):465–80.

60. Craven MP, Groom MJ. Computer games for user engagement in Attention Deficit Hyperactivity Disorder (ADHD) monitoring and therapy. In: 2015 International Conference on Interactive Technologies and Games: IEEE; 2015. p. 34–40.

61. Brooks BL, Sherman EM. Computerized neuropsychological testing to rapidly evaluate cognition in pediatric patients with neurologic disorders. Journal of Child Neurology 2012;27(8):982–91.

62. Nielsen JD, Madsen KH, Puonti O, Siebner HR, Bauer C, Madsen CG, et al. Automatic skull segmentation from MR images for realistic volume conductor models of the head: Assessment of the state-of-the-art. Neuroimage 2018;174:587–98.

63. Thielscher A, Antunes A, Saturnino GB. Field modeling for transcranial magnetic stimulation: a useful tool to understand the physiological effects of TMS? In: 2015 37th annual international conference of the IEEE engineering in medicine and biology society (EMBC): IEEE; 2015. p. 222–5.

64. Saturnino GB, Puonti O, Nielsen JD, Antonenko D, Madsen KH, Thielscher A. SimNIBS 2.1: a comprehensive pipeline for individualized electric field modelling for transcranial brain stimulation. Brain Human Body Modeling 2019:3–25.

65. Okamoto M, Dan H, Sakamoto K, Takeo K, Shimizu K, Kohno S, et al. Three-dimensional probabilistic anatomical cranio-cerebral correlation via the international 10–20 system oriented for transcranial functional brain mapping. Neuroimage 2004;21(1):99–111.

66. Caulfield KA, George MS. Optimized APPS-tDCS electrode position, size, and distance doubles the on-target stimulation magnitude in 3000 electric field models. Scientific Reports 2022;12(1):1–15.

67. Geuzaine C, Remacle JF. Gmsh: A 3-D finite element mesh generator with built-in pre- and post-processing facilities. International journal for numerical methods in engineering 2009;79(11):1309–31.

68. La Corte E, Eldahaby D, Greco E, Aquino D, Bertolini G, Levi V, et al. The frontal aslant tract: a systematic review for neurosurgical applications. Frontiers in Neurology 2021;12:641586.

69. Singh M, Fuelscher I, He J, Anderson V, Silk TJ, Hyde C. Inter-individual performance differences in the stop-signal task are associated with fibre-specific microstructure of the fronto-basal-ganglia circuit in healthy children. Cortex 2021;142:283–95.

70. Singh M, Skippen P, He J, Thomson P, Fuelscher I, Caeyenberghs K, et al. Longitudinal developmental trajectories of inhibition and white-matter maturation of the fronto-basal-ganglia circuits. Developmental Cognitive Neuroscience 2022;58:101171.

71. Bubb EJ, Metzler-Baddeley C, Aggleton JP. The cingulum bundle: anatomy, function, and dysfunction. Neuroscience & Biobehavioral Reviews 2018;92:104–27.

72. Vanderwal T, Kelly C, Eilbott J, Mayes LC, Castellanos FX. Inscapes: A movie paradigm to improve compliance in functional magnetic resonance imaging. Neuroimage 2015;122:222–32.

73. Tadel F, Baillet S, Mosher JC, Pantazis D, Leahy RM. Brainstorm: a user-friendly application for MEG/EEG analysis. Computational intelligence and neuroscience 2011;2011:1–13.

74. Sanchez CE, Richards JE, Almli CR. Age-specific MRI templates for pediatric neuroimaging. Developmental neuropsychology 2012;37(5):379–99.

75. Gaser C, Dahnke R, Thompson PM, Kurth F, Luders E. CAT-a computational anatomy toolbox for the analysis of structural MRI data. BioRxiv 2022:2022.06. 11.495736.

76. Fuchs M, Kastner J, Wagner M, Hawes S, Ebersole JS. A standardized boundary element method volume conductor model. Clinical neurophysiology 2002;113(5):702–12.

77. Dale AM, Liu AK, Fischl BR, Buckner RL, Belliveau JW, Lewine JD, et al. Dynamic statistical parametric mapping: combining fMRI and MEG for high-resolution imaging of cortical activity. neuron 2000;26(1):55–67.

78. Desikan RS, Ségonne F, Fischl B, Quinn BT, Dickerson BC, Blacker D, et al. An automated labeling system for subdividing the human cerebral cortex on MRI scans into gyral based regions of interest. 2006;31(3):968–80.

79. Hilbert D. Grundzüge einer allgemeinen Theorie der linearen Integralgleichungen. BG Teubner; 1912.

80. Zalesky A, Fornito A, Bullmore ET. Network-based statistic: identifying differences in brain networks. Neuroimage 2010;53(4):1197–207.

81. Whelan R. Effective analysis of reaction time data. The psychological record 2008;58:475–82.

82. Verbruggen F, Logan GD. Models of response inhibition in the stop-signal and stop-change paradigms. Neuroscience and Biobehavioral Reviews 2009;33(5):647–61.

83. Verbruggen F, Aron AR, Band GP, Beste C, Bissett PG, Brockett AT, et al. A consensus guide to capturing the ability to inhibit actions and impulsive behaviors in the stop-signal task. elife 2019;8:e46323.

84. Band GP, Van Der Molen MW, Logan GD. Horse-race model simulations of the stop-signal procedure. Acta psychologica 2003;112(2):105–42.

85. Lo S, Andrews S. To transform or not to transform: Using generalized linear mixed models to analyse reaction time data. Frontiers in psychology 2015;6:1171.

86. Grubbs FE. Procedures for detecting outlying observations in samples. Technometrics 1969;11(1):1–21.

87. Bender R, Lange S. Adjusting for multiple testing—when and how? Journal of clinical epidemiology 2001;54(4):343–9.

88. Konrad K, Neufang S, Thiel CM, Specht K, Hanisch C, Fan J, et al. Development of attentional networks: an fMRI study with children and adults. Neuroimage 2005;28(2):429–39.

89. Cunillera T, Brignani D, Cucurell D, Fuentemilla L, Miniussi C. The right inferior frontal cortex in response inhibition: A tDCS–ERP co-registration study. Neuroimage 2016;140:66–75.

90. Castro-Meneses LJ, Johnson BW, Sowman PF. Vocal response inhibition is enhanced by anodal tDCS over the right prefrontal cortex. Experimental Brain Research 2016;234(1):185–95.

91. Jacobson L, Javitt DC, Lavidor M. Activation of inhibition: diminishing impulsive behavior by direct current stimulation over the inferior frontal gyrus. Journal of Cognitive Neuroscience 2011;23(11):3380–7.

92. Stramaccia DF, Penolazzi B, Sartori G, Braga M, Mondini S, Galfano G. Assessing the effects of tDCS over a delayed response inhibition task by targeting the right inferior frontal gyrus and right dorsolateral prefrontal cortex. Experimental Brain Research 2015;233(8):2283–90.

93. Friehs MA, Frings C. Cathodal tDCS increases stop-signal reaction time. Cognitive, Affective, Behavioral Neuroscience 2019;19(5):1129–42.

94. Friehs MA, Frings C. Pimping inhibition: Anodal tDCS enhances stop-signal reaction time. Journal of Experimental Psychology: Human Perception and Performance 2018;44(12):1933.

95. Friehs MA, Brauner L, Frings C. Dual-tDCS over the right prefrontal cortex does not modulate stop-signal task performance. Experimental brain research 2021;239:811–20.

96. London RE, Slagter HA. No effect of transcranial direct current stimulation over left dorsolateral prefrontal cortex on temporal attention. Journal of cognitive neuroscience 2021;33(4):756–68.

97. Rushby JA, De Blasio FM, Logan JA, Wearne T, Kornfeld E, Wilson EJ, et al. tDCS effects on task-related activation and working memory performance in traumatic brain injury: A within group randomized controlled trial. Neuropsychological Rehabilitation 2020:1–23.

98. Kang E-K, Kim D-Y, Paik N-JJJorm. Transcranial direct current stimulation of the left prefrontal cortex improves attention in patients with traumatic brain injury: a pilot study. 2012;44(4):346–50.

99. O’Neil-Pirozzi TM, Doruk D, Thomson JM, Fregni FJIJoN. Immediate memory and electrophysiologic effects of prefrontal cortex transcranial direct current stimulation on neurotypical individuals and individuals with chronic traumatic brain injury: a pilot study. 2017;127(7):592–600.

100. Minarik T, Berger B, Althaus L, Bader V, Biebl B, Brotzeller F, et al. The importance of sample size for reproducibility of tDCS effects. Frontiers in human neuroscience 2016;10:453.

101. Lesniak M, Polanowska K, Seniów J, Czlonkowska A. Effects of repeated anodal tDCS coupled with cognitive training for patients with severe traumatic brain injury: a pilot randomized controlled trial. The Journal of head trauma rehabilitation 2014;29(3):E20–E9.

102. Filmer HL, Varghese E, Hawkins GE, Mattingley JB, Dux PE. Improvements in attention and decision-making following combined behavioral training and brain stimulation. Cerebral Cortex 2017;27(7):3675–82.

103. Allenby C, Falcone M, Bernardo L, Wileyto EP, Rostain A, Ramsay JR, et al. Transcranial direct current brain stimulation decreases impulsivity in ADHD. Brain Stimulation 2018;11(5):974–81.

104. Monte-Silva K, Kuo M-F, Liebetanz D, Paulus W, Nitsche MA. Shaping the optimal repetition interval for cathodal transcranial direct current stimulation (tDCS). Journal of neurophysiology 2010;103(4):1735–40.

105. Alonzo A, Brassil J, Taylor JL, Martin D, Loo CK. Daily transcranial direct current stimulation (tDCS) leads to greater increases in cortical excitability than second daily transcranial direct current stimulation. Brain Stimulation 2012;5(3):208–13.

106. Stein A, Iyer KK, Barlow KM. Cerebral activation of attention and working memory in traumatic brain injury. In: Diagnosis and Treatment of Traumatic Brain Injury: Elsevier; 2022. p. 151–67.

107. Gu S, Satterthwaite TD, Medaglia JD, Yang M, Gur RE, Gur RC, et al. Emergence of system roles in normative neurodevelopment. Proceedings of the National Academy of Sciences 2015;112(44):13681–6.

108. Satterthwaite TD, Wolf DH, Erus G, Ruparel K, Elliott MA, Gennatas ED, et al. Functional maturation of the executive system during adolescence. Journal of Neuroscience 2013;33(41):16249–61.

109. Cahoon GD, Davison TE. Prediction of compliance with MRI procedures among children of ages 3 years to 12 years. Pediatric Radiology 2014;44(10):1302–9.

110. Castellanos NP, Paúl N, Ordóñez VE, Demuynck O, Bajo R, Campo P, et al. Reorganization of functional connectivity as a correlate of cognitive recovery in acquired brain injury. Brain 2010;133(8):2365–81.

111. Huang M-X, Theilmann RJ, Robb A, Angeles A, Nichols S, Drake A, et al. Integrated imaging approach with MEG and DTI to detect mild traumatic brain injury in military and civilian patients. Journal of neurotrauma 2009;26(8):1213–26.

112. Lewine JD, Davis JT, Sloan JH, Kodituwakku P, Orrison Jr WW. Neuromagnetic assessment of pathophysiologic brain activity induced by minor head trauma. American Journal of Neuroradiology 1999;20(5):857–66.

113. Alhourani A, Wozny TA, Krishnaswamy D, Pathak S, Walls SA, Ghuman AS, et al. Magnetoencephalography-based identification of functional connectivity network disruption following mild traumatic brain injury. Journal of neurophysiology 2016;116(4):1840–7.

114. Robb Swan A, Nichols S, Drake A, Angeles A, Diwakar M, Song T, et al. Magnetoencephalography slow-wave detection in patients with mild traumatic brain injury and ongoing symptoms correlated with long-term neuropsychological outcome. Journal of neurotrauma 2015;32(19):1510–21.

115. Mo J, Liu Y, Huang H, Ding M. Coupling between visual alpha oscillations and default mode activity. Neuroimage 2013;68:112–8.

116. Knyazev GG, Slobodskoj-Plusnin JY, Bocharov AV, Pylkova LV. The default mode network and EEG alpha oscillations: an independent component analysis. Brain research 2011;1402:67–79.

117. Jann K, Dierks T, Boesch C, Kottlow M, Strik W, Koenig T. BOLD correlates of EEG alpha phase-locking and the fMRI default mode network. Neuroimage 2009;45(3):903–16.

118. Samogin J, Liu Q, Marino M, Wenderoth N, Mantini D. Shared and connection-specific intrinsic interactions in the default mode network. Neuroimage 2019;200:474–81.

119. Clancy KJ, Andrzejewski JA, You Y, Rosenberg JT, Ding M, Li W. Transcranial stimulation of alpha oscillations up-regulates the default mode network. Proceedings of the National Academy of Sciences 2022;119(1):e2110868119.

120. Kaltiainen H, Liljeström M, Helle L, Salo A, Hietanen M, Renvall H, et al. Mild traumatic brain injury affects cognitive processing and modifies oscillatory brain activity during attentional tasks. Journal of neurotrauma 2019;36(14):2222–32.

121. Popescu M, Hughes JD, Popescu E-A, Riedy G, DeGraba TJ. Reduced prefrontal MEG alpha-band power in mild traumatic brain injury with associated posttraumatic stress disorder symptoms. Clinical neurophysiology 2016;127(9):3075–85.

122. Palacios EM, Sala-Llonch R, Junque C, Roig T, Tormos JM, Bargallo N, et al. Resting-state functional magnetic resonance imaging activity and connectivity and cognitive outcome in traumatic brain injury. JAMA Neurol 2013;70(7):845–51.

123. Stein A, Iyer KK, Khetani AM, Barlow KM. Changes in working memory-related cortical responses following pediatric mild traumatic brain injury: A longitudinal fMRI study. Journal of Concussion 2021;5:20597002211006541.

124. Leech R, Sharp DJ. The role of the posterior cingulate cortex in cognition and disease. Brain 2014;137(1):12–32.

125. Straume-Naesheim T, Andersen T, Bahr R. Reproducibility of computer based neuropsychological testing among Norwegian elite football players. British journal of sports medicine 2005;39(suppl 1):i64–i9.

126. Collie A, Maruff P, Makdissi M, McStephen M, Darby DG, McCrory P. Statistical procedures for determining the extent of cognitive change following concussion. British Journal of Sports Medicine 2004;38(3):273–8.

127. Tseng P, Hsu T-Y, Chang C-F, Tzeng OJ, Hung DL, Muggleton NG, et al. Unleashing potential: transcranial direct current stimulation over the right posterior parietal cortex improves change detection in low-performing individuals. Journal of Neuroscience 2012;32(31):10554–61.

128. Li LM, Uehara K, Hanakawa T. The contribution of interindividual factors to variability of response in transcranial direct current stimulation studies. Frontiers in Cellular Neuroscience 2015;9:181.

129. Meng Q, Zhu Y, Yuan Y, Liu J, Ye L, Kong W, et al. A Novel Approach to Modulating Response Inhibition: Multi-Channel Beta Transcranial Alternating Current Stimulation. Asian Journal of Psychiatry 2023:103872.

130. Arciniega H, Gözenman F, Jones KT, Stephens JA, Berryhill ME. Frontoparietal tDCS benefits visual working memory in older adults with low working memory capacity. Frontiers in aging neuroscience 2018;10:57.

131. Gözenman F, Berryhill ME. Working memory capacity differentially influences responses to tDCS and HD-tDCS in a retro-cue task. Neuroscience letters 2016;629:105–9.

132. Datta A, Truong D, Minhas P, Parra LC, Bikson M. Inter-individual variation during transcranial direct current stimulation and normalization of dose using MRI-derived computational models. Frontiers in psychiatry 2012;3:91.

133. Bikson M, Rahman A, Datta A. Computational models of transcranial direct current stimulation. Clinical EEG and neuroscience 2012;43(3):176–83.

134. Li LM, Violante IR, Leech R, Hampshire A, Opitz A, McArthur D, et al. Cognitive enhancement with Salience Network electrical stimulation is influenced by network structural connectivity. Neuroimage 2019;185:425–33.

135. Cai Y, Li S, Liu J, Li D, Feng Z, Wang Q, et al. The role of the frontal and parietal cortex in proactive and reactive inhibitory control: a transcranial direct current stimulation study. Journal of cognitive neuroscience 2016;28(1):177–86.

136. Alekseichuk I, Wischnewski M, Opitz A. A minimum effective dose for (transcranial) alternating current stimulation. Brain Stimulation: Basic, Translational, and Clinical Research in Neuromodulation 2022;15(5):1221–2.

137. Caulfield KA, Indahlastari A, Nissim NR, Lopez JW, Fleischmann HH, Woods AJ, et al. Electric field strength from prefrontal transcranial direct current stimulation determines degree of working memory response: a potential application of reverse-calculation modeling? Neuromodulation: Technology at the Neural Interface 2022.

138. Hoy KE, Emonson MR, Arnold SL, Thomson RH, Daskalakis ZJ, Fitzgerald PB. Testing the limits: investigating the effect of tDCS dose on working memory enhancement in healthy controls. Neuropsychologia 2013;51(9):1777–84.

139. Evans C, Bachmann C, Lee JS, Gregoriou E, Ward N, Bestmann S. Dose-controlled tDCS reduces electric field intensity variability at a cortical target site. Brain stimulation 2020;13(1):125–36.

140. McAllister TW, Sparling MB, Flashman LA, Saykin AJ. Neuroimaging findings in mild traumatic brain injury. Journal of clinical and experimental neuropsychology 2001;23(6):775–91.

141. Carlson HL, Giuffre A, Ciechanski P, Kirton A. Electric field simulations of transcranial direct current stimulation in children with perinatal stroke. Frontiers in Human Neuroscience 2023;17:1075741.

142. Buchanan DM, Bogdanowicz T, Khanna N, Lockman-Dufour G, Robaey P, D’Angiulli A. Systematic review on the safety and tolerability of transcranial direct current stimulation in children and adolescents. Brain sciences 2021;11(2):212.

143. Sierawska A, Splittgerber M, Moliadze V, Siniatchkin M, Buyx A. Transcranial Direct Current Stimulation (tDCS) in Pediatric Populations—–Voices from Typically Developing Children and Adolescents and their Parents. Neuroethics 2023;16(1):3.

144. Andrade AC, Magnavita GM, Allegro JVBN, Neto CEBP, Lucena RdCS, Fregni F. Feasibility of transcranial direct current stimulation use in children aged 5 to 12 years. Journal of child neurology 2014;29(10):1360–5.

145. Kirton A, Ciechanski P, Zewdie E, Andersen J, Nettel-Aguirre A, Carlson H, et al. Transcranial direct current stimulation for children with perinatal stroke and hemiparesis. Neurology 2017;88(3):259–67.

146. Pilloni G, Choi C, Shaw MT, Coghe G, Krupp L, Moffat M, et al. Walking in multiple sclerosis improves with tDCS: a randomized, double-blind, sham-controlled study. Annals of Clinical and Translational Neurology 2020;7(11):2310–9.

147. Sale MV, Ridding MC, Nordstrom MAJJoN. Cortisol inhibits neuroplasticity induction in human motor cortex. 2008;28(33):8285–93.

148. Zelazo PD, Anderson JE, Richler J, Wallner-Allen K, Beaumont JL, Conway KP, et al. NIH Toolbox Cognition Battery (CB): validation of executive function measures in adults. Journal of the International Neuropsychological Society 2014;20(6):620.

