## Supplementary for "The effect of a single session of tDCS on attention in pediatric acquired brain injury: Characterising inter-individual structural and functional network response variability"

### Supplementary Methods

#### Definition of TBI severity and tDCS contraindications

TBI injury severity was defined according to the Glasgow Coma Scale (GCS) where mild was considered to be a score of between 13 and 15 upon ED evaluation and <24 h post-traumatic amnesia (PTA); moderate was considered as a loss of consciousness (LOC) of between 30 min and 24 hours, GCS between 9 and 12 and PTA between 24 h and 7 days; and severe was considered as GCS<9 and PTA for more than 7 days.[1]

Contraindications to tDCS included metal implants, serious or unstable illness or medical condition, current or suspected pregnancy or current lactation, skull defects underneath stimulation sites, scalp wound/skin problem preventing placement of HD-EEG and/or stimulation leads, and epilepsy or other seizure disorders.

#### Clinical, neurocognitive and behavioural questionnaires

*CNS Vital Signs*

CNS Vital Signs (CNSVS) was used to provide information on cognition. CNSVS is a computerised assessment procedure that utilizes scientifically validated, objective, and reliable computerized neuropsychological tests to evaluate the neurocognitive status of patients and covers a range of mental processes from simple motor performance, attention, memory, to executive functions. It provides a rapid assessment of cognition (30 minutes) and is well tolerated and valid in paediatric neurology patients.[2]

*BRIEF2*

The parent version of the Behaviour Rating Inventory for Executive Functioning Second Edition was used to assess children's executive functioning and likelihood of ADHD. The BRIEF2[3] is a questionnaire validated for use in pediatric TBI populations.[4] It consists of eight clinical scales (Inhibit, Shift, Emotional Control, Initiate, Working Memory, Plan/Organize, Organization of Materials, Monitor) and two validity scales (Inconsistency and Negativity).

#### Outcome measures

*Flanker task*

The flanker task[5] is an inhibition task requiring participants to respond to a centrally-presented target stimulus (an arrow facing one direction) while ignoring flanking task-irrelevant information (arrows facing the opposite direction of the target arrow).[5] The NIH Toolbox flanker task (iPad) was used in this study.[6] This toolbox consists of a battery of cognitive tests designed to be used in clinical populations such as ABI. Participants performed the test seated with the iPad on the desk in front of them and were instructed to use the index finger of their dominant hand to respond. The duration of the task was approximately 3 minutes, consisting of a mixture of 24 congruent and incongruent trials.

*Stop signal Task*

As a secondary outcome, gamified Stop Signal RT[7] was also assessed immediately prior to and after tDCS. The Stop Signal Task[8] is used to measure response inhibition for prepotent responses. During this task, participants must respond as quickly as possible to a target stimulus but inhibit their response when the stop signal is displayed. Participants completed the SST on a Dell Latitude 5411 laptop (Dell Inc., Round Rock, Texas) placed on a desk, while seated in a comfortable chair. Participants used their dominant hand to press the arrow keys on the keyboard. A gamified version of the Stop-Signal Task (known as the Stop Signal Game or 'SSG') was used in this study. The duration of the SSG was approximately 12 minutes (two blocks of 6 mins separated by a 15 second break) involving a total of 200 trials (including 104 stop trials). In this game, the participant must find their way out of an enchanted forest by pressing the left or right arrow keys as quickly as possible (as instructed by a magical fairy). However, the participant must withhold their response if they hear a beep from an evil witch (auditory stop signal), or risk being 'lured' further into the forest. The gamified SST used in this study replicates a more natural setting compared to the traditional SST, thus eliciting more natural responses without sacrificing experimental control. The gamified SST has been validated against the traditional SST using healthy populations, where comparable performance was found between both tasks.[7] The Stop Signal Game[7] used was piloted in 10 healthy children prior to use in the study, and valid responses were obtained.

#### Attention training during tDCS

During tDCS stimulation (active and sham), participants completed attention training tasks. Gamified tasks were chosen to enhance participant engagement.[9-11] A gamified Go/No-Go task, 'Wormy Fruit', was presented first to participants for eight minutes.[12] The Go/No-Go task is an inhibitory control task requiring participants to respond to certain target stimuli while inhibiting their response to non-target stimuli.[13] Wormy Fruit was started simultaneously with stimulation and lasted eight minutes. After two-minutes of rest, participants engaged in a gamified continuous performance task (CPT), called 'SnappyApp' (eight minutes).[12] The CPT measures sustained attention, vigilance and impulsivity.[14]

*Wormy Fruit Go/No-Go Task*

The Go/No-Go Task[15] is an inhibition task which requires participants to provide a response to a "go" stimulus whilst refraining from reacting to a "no-go" stimulus. The Go/No-Go task is sensitive to participant performance in child ABI populations[16,17], where children with TBI have been shown to have more commission errors (failure to withhold a response on a "no-go" trial) compared to uninjured children.[18] In this study, a gamified version of the Go/No-Go task known as 'Wormy Fruit' was used.[12] In this task, participants were presented with either a "Go" stimulus (whole red apple), or "No-Go" stimulus (red apple with a worm inside). Participants were required to 'eat' the apple (by pressing spacebar) providing it is whole (without a worm) or withhold their response if the apple contains a worm. Participants were seated in a comfortable chair and completed the task on a Dell Latitude 5411 (Dell Technologies; Round Rock, Texas) 14-inch laptop placed directly in front of them on a desk using their dominant hand. Participant reaction time, omission errors and commission errors were recorded. Stimulus presentation duration was 200 ms, the interstimulus interval was 200ms and the task duration was eight minutes, allowing for a total of 240 stimulus presentations (where 179 (~75%) were 'Go' stimuli, and 61 (~25%) were 'No-Go' stimuli) (presented pseudo-randomly). As this task had not previously been used in a pediatric population, 'Wormy Fruit' was piloted in 10 healthy children to ensure validity in pediatric populations, where valid responses were obtained.

*'SnappyApp' Continuous Performance Test*

The traditional Conner's Continuous Performance Test (CPT)[14] requires participants to focus their attention for an extended period of time to detect a target stimulus. The CPT measures inattentiveness, impulsivity, sustained attention and vigilance. In this study, a gamified version of the CPT known as 'SnappyApp'[12] was used. In this task, participants were briefly and continuously shown pictures of different fruit and had to only react to the target stimulus (bananas which appeared directly after cherries) by pressing spacebar. Participants were seated in a comfortable chair and completed the task on a Dell Latitude 5411 (Dell Technologies; Round Rock, Texas) 14-inch laptop placed directly in front of them on a desk using their dominant hand. Participant reaction time, omission and commission errors were recorded. Stimulus presentation duration was 200 ms, the interstimulus interval was 200ms and the total task duration was eight minutes, resulting in a total of 240 stimulus presentations (where 24 (10%) were target stimuli, and 216 (90%) were non-target stimuli). As this task had not been used in a pediatric population, 'SnappyApp' was piloted in 10 healthy children to ensure that the task produced valid results in a pediatric population., where valid responses were obtained.

*Sensations and adverse effects questionnaire*

#### Questionnaire of sensations related to transcranial direct current stimulation (tDCS)

*(To be filled in by participants)*

Investigator: ______________________________

Participant RND number:______________________________Date:___/____/____

Session number: ______________________________

Number of previous stimulations ________________

tDCS Intensity: ________ mA

Electrode dimensions: anode: _____x_____ cathode _____x_____ shape _______

Did you experience any discomfort during the electrical stimulation? Please indicate the degree of intensity of your discomfort according to the following scale:

- None = I did not feel this sensation
- Mild = I mildly felt this sensation
- Moderate = I felt this sensation
- Strong = I felt this sensation to a considerable degree

| In the first stimulation block I felt (to be filled in by participant, if it is possible please write where you felt the sensation): | | | | |
|---|---|---|---|---|
| | None | Mild | Moderate | Strong |
| Itching | ☐ | ☐ | ☐ | ☐ |
| Pain | ☐ | ☐ | ☐ | ☐ |
| Burning | ☐ | ☐ | ☐ | ☐ |
| Warmth/Heat | ☐ | ☐ | ☐ | ☐ |
| Metallic/Iron taste | ☐ | ☐ | ☐ | ☐ |
| Fatigue | ☐ | ☐ | ☐ | ☐ |
| Decreased alertness | ☐ | ☐ | ☐ | ☐ |
| Other | ☐ | ☐ | ☐ | ☐ |

Duration (multiple options allowed)

☐ Only initially ☐ It stopped in the middle of the session ☐ It stopped at the end of the session

How much did these sensations affect you?

☐ Not at all ☐ Slightly ☐ Considerably ☐ Much ☐ Very much

In this stimulation session, do you believe that you received a real or placebo stimulation?

☐ real ☐ sham ☐ I don't know

Additional comments: ______________________________________________

Signature of Investigator: ______________________

Date: ______________________

*Supplementary Figure 1: Sensations and adverse effects questionnaire used at the end of each tDCS session. The questionnaire was delivered in the form of an interview. Adapted from Fertonani et al. 2010*[19]

**Functional connectivity**

*HD-EEG pre-processing*

First, the HD-EEG data were trimmed to remove the first and last 60 seconds of recording as participants showed greater restlessness during these periods, leaving three minutes of continuous EEG data. Next, we opted to remove 19 peripheral channels using the 'pop_select' function as described in Angelini et al. 2016[20] and Calbi et al. 2019[21], leaving a total of 110 channels in analysis. A high-pass filter was applied at 0.5 Hz and the channels were re-referenced to the average montage using the 'pop_reref' function. Independent component analysis (ICA) was conducted using the 'runica (fastica)' function labelling independent components (ICs) with the 'pop_iclabel' function. Using the 'select_IC' and 'pop_subcomp' functions, ICs identified as muscle, eye, heart, line noise, channel noise or 'other' with greater than 50% probability were removed. The data was re-referenced to average, followed by rejection of bad channels using the 'pop_rejchan' function with a threshold of 3. EEGs were then spherically interpolated from neighbouring channels using the 'pop_interp' function. An average of 7 channels were interpolated, predominantly from parieto-occipital regions. Bandpass filtering was then conducted using a 4$^{th}$ order Butterworth filter between 0.5 – 45 Hz. Lastly, data was epoched into 6 segments of 30 s each, as small epochs are associated with greater stabilization of network topology.[22]

*Spectral power*

Mean spectral power at the sensor level was computed via Welch's estimate.[23] Mean spectral power was extracted all channels, as well as in a subgroup of a frontal division, temporal division and parieto-occipital division of channels.[21]

Mean spectral power across the whole brain as well as a subgroup of a frontal division, temporal division and parieto-occipital division of channels was compared between HC and ABI participants across delta, theta, alpha, beta, gamma, and broadband frequencies in the resting EO and EC conditions using two sample t-tests.

*ROI definitions*

Connectivity values were extracted from the nodes defined in each ROI and averaged to give a mean connectivity value in each ROI. DMN nodes were defined according to the Desikan Killiany atlas bilateral 'isthmus cingulate', 'posterior cingulate', 'precuneus', 'rostral anterior cingulate', 'parahippocampal' and 'rostral middle frontal' nodes.[24] SN nodes were defined as the bilateral 'caudal anterior cingulate' and 'insula' nodes.[25,26] ECN nodes were defined as the left 'middle temporal', bilateral 'superior parietal', bilateral 'caudal middle frontal', and right caudal anterior cingulate'.[27]

#### Structural connectivity

*MRI acquisition*

T1 scans were acquired using MPRAGE, TR 1880.0ms, TE 2.32 ms, voxel size=0.9x0.9x0.9mm, 192 slices. T2 scans were acquired using SPC FLAIR, TR 5000 ms, TE 389 ms, voxel size 0.9 x 0.9 x0.9 mm, 192 slices. For diffusion weighted imaging (DWI), three diffusion scans were collected in short blocks (< 2.5 mins), including 8xb=0, 30xb=900 s/mm$^2$, 30xb=2000 s/mm$^2$.

*DTI preprocessing and model fitting*

All steps described below were completed in MRtrix3Tissue (version 5.2.8), a fork of the MRtrix3 software platform.[28] This pipeline was conducted using DWI data in n=10 participants acquired using the following acquisition scheme: b = 900 s/mm$^2$ (30 directions, b0 = 1), as this was the highest b value with the most consistent number of directions across all subjects. Raw DWI images for each participant underwent preprocessing using the recommended pipeline in MRtrix3. Briefly, this included denoising[29], Gibbs unringing [30], and correction for motion, eddy current, and bias field inhomogeneity related artefacts[31,32].

Following preprocessing, tissue-specific response functions for white matter (WM), grey matter (GM) and cerebrospinal fluid (CSF) were estimated for each subject using an unsupervised method and averaged to generate group-level values[33] The data was then up-sampled to a voxel size of 1.5mm$^3$ and single-shell 3-tissue Constrained Spherical Deconvolution (SS3T-CSD) was conducted to obtain Fibre Orientation functions (FODfs) within WM[34]. A CSD-based approach was chosen as the primary model-fitting paradigm due to its ability to accurately reconstruct diffusion behaviour within voxels containing complex fibre orientations, which in some estimates range from 60-90% of all WM voxels within the brain.[35] Individual subject FODfs were then intensity normalized [36] and averaged to form a study-specific population template using affine linear registration. Subsequently, the template image was warped to MNI space and estimates of fibre density (FD), a measure of intravoxel microstructural integrity [37], was computed from the whole brain fixel maps for each subject. Fixels refer to fibre-specific populations within a voxel, allowing for the calculation of metrics that are arguably more biologically plausible compared to other voxelwise approaches [38]. In addition to CSD, we also conducted traditional DTI based modelling. Here, each participant's pre-processed image was fit with a diffusion tensor model using the "dwi2tensor" command to generate tensor maps of diffusion behaviour across the whole brain. The application of the tensor model resulted in whole brain voxelwise fractional anisotropy (FA) maps for each individual participant. The FA maps were then registered to MNI space using the warps previously generated from the FODf registration.

*Tractography and outcome measures*

Drawing from the work of Li et al., 2019[39], tracts of interest underlying the salience (SN) and default mode (DMN) networks were used to examine associations between structural connectivity and RT change following tDCS. These included the frontal aslant tract (FAT), a structural network connecting the inferior frontal gyri/anterior insula with the presupplementary motor area[40] and the cingulum bundle (CG), a prominent WM tract connecting frontal, parietal and temporal cortices[41]. To delineate the FAT in each hemisphere, manual probabilistic tractography was applied to the population template following the approach done in previous diffusion MRI studies in typically developing children[42,43] The CG was delineated in the population template using TractSeg, an automated tool that provides study-specific segmentations of 72 major WM tracts using the peaks of the FODfs as input.[44] Supplementary Figure 1 visualizes each tract of interest used in the current study. Lastly, to examine associations between baseline structural connectivity within the SN and DMN and RT change post-tDCS, mean FD and FA values for each participant was extracted from all tracts of interest for subsequent correlation analysis.

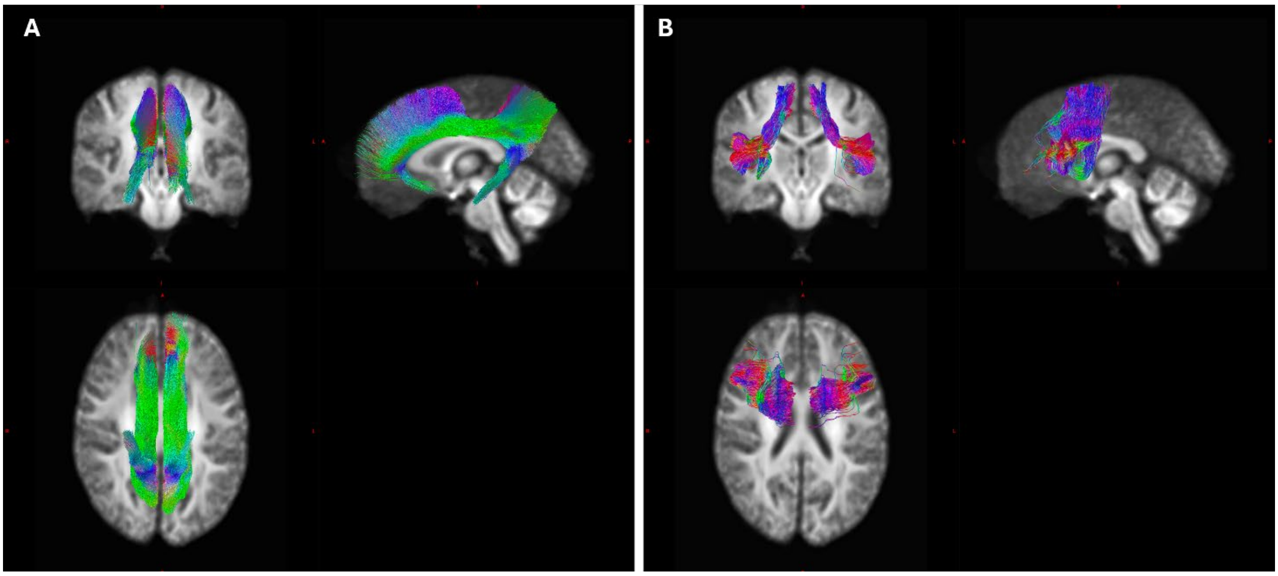


*Supplementary Figure 2. Tractograms overlaid on the subject-specific population template. (A) Left and right cingulate gyrus (CG); (B) left and right frontal aslant tract (FAT)*

**LMM equations: post-tDCS flanker RT**

$$[\text{post tDCS RT}]_{ij} = \gamma_{00} + u_{0j} + \gamma_{10}[\text{pre tDCS RT}]_{ij} + \gamma_{20}[\text{dlPFC tDCS}]_{ij} + \gamma_{30}[\text{IFG tDCS}]_{ij} + e_{ij}$$

Supplementary Equation 1: Linear mixed model for post-tDCS RT across tDCS arms.
$i$ = time point, $j$ = participant, $\gamma$ = mean estimate, $u$ = random effect for participant, and $e$ = residual error
tDCS, transcranial direct current stimulation; RT, reaction time; dlPFC, dorsolateral prefrontal cortex; IFG, inferior frontal gyrus

$$[\text{post tDCS RT}]_{ij} = \gamma_{00} + u_{0j} + \gamma_{10}[\text{pre tDCS RT}]_{ij} + \gamma_{20}[\text{dlPFC tDCS}]_{ij} + \gamma_{30}[\text{IFG tDCS}]_{ij} + \gamma_{40}[\text{session order}]_{ij} + e_{ij}$$

Supplementary Equation 2: Linear mixed model for post-tDCS RT across tDCS arms including the effect of session order.
$i$ = time point, $j$ = participant, $\gamma$ = mean estimate, $u$ = random effect for participant, and $e$ = residual error
tDCS, transcranial direct current stimulation; RT, reaction time; dlPFC, dorsolateral prefrontal cortex; IFG, inferior frontal gyrus

### Supplementary Results

#### Reaction time

Supplementary Table 1: Mean baseline RT across sessions

| | ABI Flanker RT (s, 95% CI) | ABI SSRT (s, 95% CI) | HC Flanker RT (s, 95% CI) | HC SSRT (s, 95% CI) |
|---|---|---|---|---|
| **dlPFC** | 0.70 (0.50-0.89) | 0.44 (0.38-0.49) | 0.59 (0.47-0.71) | 0.47 (0.39-0.55) |
| **IFG** | 0.71 (0.52-0.90) | 0.43 (0.39-0.47) | 0.61 (0.50-0.72) | 0.46 (0.37-0.55) |
| **sham** | 0.57 (0.48-0.65) | 0.43 (0.39-0.48) | 0.56 (0.47-0.65) | 0.47 (0.35-0.59) |

ABI, acquired brain injury; HC, healthy control; SD, standard deviation; dlPFC, dorsolateral prefrontal cortex; IFG, inferior frontal gyrus; s, seconds; CI, confidence interval

#### Post-tDCS RT across tDCS arms

Supplementary Table 2: Mean RT change across sessions

| | ABI Flanker RT change (s, 95% CI) | ABI SSRT change (s, 95% CI) | HC Flanker RT change (s, 95% CI) | HC SSRT change (s, 95% CI) |
|---|---|---|---|---|
| **dlPFC** | -0.08 (-0.19, 0.02) | 0.03 (-0.03, 0.10) | -0.04 (-0.09, 0.01) | 0.04 (-0.09, 0.16) |
| **IFG** | -0.07 (-0.21, 0.08) | 0.03 (-0.04, 0.10) | -0.04 (-0.12, 0.05) | 0.03 (-0.03, 0.09) |
| **sham** | 0.05 (-0.04, 0.14) | 0.00 (-0.04, 0.04) | 0.01 (-0.03, 0.05) | -0.00 (-0.06, 0.06) |

ABI, acquired brain injury; HC, healthy control; SD, standard deviation; dlPFC, dorsolateral prefrontal cortex; IFG, inferior frontal gyrus; s, seconds; CI, confidence interval

Supplementary Table 3: Model summary of primary outcome LMM in ABI participants

**ANOVA Summary**

| Effect | df | F | p |
|---|---|---|---|
| Flanker_pre_RT | 1, 1064.34 | 25.108 | < .001 |
| Region | 2, 1057.18 | 0.396 | 0.673 |

**Fixed Effects Estimates**

| Term | Estimate | SE | df | t | p |
|---|---|---|---|---|---|
| Intercept | 0.519 | 0.057 | 18.203 | 9.116 | < .001 |
| Flanker_pre_RT | 0.161 | 0.032 | 1064.342 | 5.011 | < .001 |
| Region (1) | -0.018 | 0.020 | 1056.856 | -0.882 | 0.378 |
| Region (2) | 0.011 | 0.020 | 1057.001 | 0.532 | 0.595 |

**Estimated Marginal Means**

| | | | 95% CI | |
|---|---|---|---|---|
| **Flanker_pre_RT** | **Estimate** | **SE** | **Lower** | **Upper** |
| 0.154 | 0.543 | 0.055 | 0.435 | 0.652 |
| 0.657 | 0.625 | 0.053 | 0.521 | 0.728 |
| 1.160 | 0.706 | 0.055 | 0.597 | 0.814 |

Supplementary Table 4: Model summary of exploratory LMM with session order in ABI participants

**ANOVA Summary**

| Effect | df | F | p |
|---|---|---|---|
| Flanker_pre_RT | 1, 1061.00 | 22.087 | < .001 |
| Region | 2, 1055.04 | 0.485 | 0.616 |
| session_order | 2, 1055.82 | 0.478 | 0.620 |

**Fixed Effects Estimates**

| Term | Estimate | SE | df | t | p |
|---|---|---|---|---|---|
| Intercept | 0.523 | 0.057 | 18.379 | 9.106 | < .001 |
| Flanker_pre_RT | 0.155 | 0.033 | 1060.998 | 4.700 | < .001 |
| Region (1) | -0.020 | 0.020 | 1054.751 | -0.977 | 0.329 |
| Region (2) | 0.012 | 0.020 | 1054.957 | 0.584 | 0.559 |
| session_order (1) | 0.019 | 0.021 | 1056.843 | 0.914 | 0.361 |
| session_order (2) | -0.015 | 0.020 | 1054.817 | -0.758 | 0.449 |

**Estimated Marginal Means**

| | | | 95% CI | |
|---|---|---|---|---|
| **Flanker_pre_RT** | **Estimate** | **SE** | **Lower** | **Upper** |
| 0.154 | 0.546 | 0.056 | 0.437 | 0.656 |
| 0.657 | 0.625 | 0.053 | 0.520 | 0.729 |
| 1.160 | 0.703 | 0.056 | 0.594 | 0.812 |

Supplementary Table 5: Number of participants included in SSRT analysis over time and across tDCS arm

| | ABI | | HC | |
|---|---|---|---|---|
| | **Pre (n)** | **Post (n)** | **Pre (n)** | **Post (n)** |
| **dlPFC** | 12 | 13 | 14 | 14 |

| | | | | |
|---|---|---|---|---|
| **IFG** | 12 | 13 | 12 | 11 |
| **sham** | 14 | 10 | 13 | 12 |

ABI, acquired brain injury; HC, healthy control; SD, standard deviation; dlPFC, dorsolateral prefrontal cortex; IFG, inferior frontal gyrus

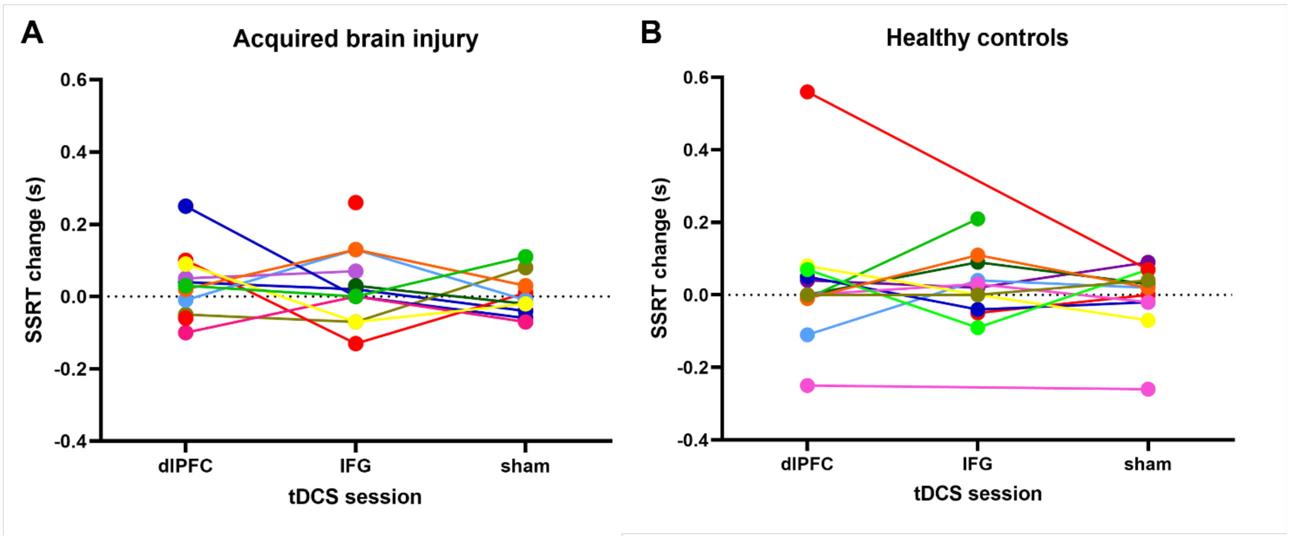


*Supplementary Figure 3: SSRT change across tDCS arm (dlPFC, IFG, sham) in ABI and HC groups. Colours represent within-subject data.*
*SSRT, stop signal reaction time; tDCS, transcranial direct current stimulation; s, seconds; dlPFC, dorsolateral prefrontal cortex; IFG, inferior frontal gyrus; ABI, acquired brain injury; HC, healthy control*

For SSRT, there was no significant effect in either the ABI or HC group for time (pre vs. post tDCS; ABI: F=1.83, p=0.19; HC: F=0.16, p=0.69) or tDCS arm (dlPFC, IFG, sham; ABI: F=0.29, p=0.75; HC: F=0.25, p=0.78; Supplementary Figure 2). Furthermore, there was no effect of group (ABI vs. HC: F=0.01, p=0.91). There were no significant differences in the number of participants in the pre and post-tDCS timepoints across tDCS arms in either the ABI or HC groups (Supplementary Table 5).

**Baseline functional connectivity differences between groups**

Supplementary Table 6: Nodes in subnetworks showing significant differences between ABI and HC participants

| EEG condition | Power spectral band | Direction of comparison | Nodes in subnetworks | T-statistic threshold | p val |
|---|---|---|---|---|---|
| RC | Alpha | Decreased in ABI | **1 subnetwork:**<br>l.cuneus<br>l.posteriorcingulate<br>r.rostralmiddlefrontal<br>r.superiorfrontal<br>r.caudalmiddlefrontal<br>r.posteriorcingulate<br>r.paracentral<br>r.inferiorparietal | 3.8 | p=0.003 |
| RC | Gamma | Increased in ABI | **1 subnetwork:**<br>r.cuneus<br>r.isthmuscingulate<br>r.paracentral<br>r.precuneus<br>l.inferiorparietal<br>l.superiorfrontal<br>l.pericalcarine | 3.1 | p=0.009 |
| RO | Delta | Increased in ABI | **2 subnetworks**<br><br>**Network 1:**<br>l.insula<br>r.isthmuscingulate<br>r.cuneus<br>r.posteriorcingulate<br><br>**Network 2:**<br>l.parsopercularis<br>l.posteriorcingulate<br>l.fusiform<br>l.transversetemporal<br>r.superiorfrontal | 3.1 | Network 1: p=0.013<br><br>Network 2: p=0.012 |

| | | | | | |
|---|---|---|---|---|---|
| RO | Theta | Increased in ABI | **1 subnetwork:**<br>l.parsopercularis<br>l.parsorbitalis<br>l.parstriangularis<br>l.inferiorparietal<br>l.posteriorcingulate<br>l.entorhinal<br>r.parstriangularis<br>r.fusiform<br>r.parstriangularis<br>r.insula<br>r.precuneus | 3.1 | p=0.016 |

NBS corrected for age, 5000 permutations, comparisons at alpha=0.05

l, left; r, right; ABI, acquired brain injury, NBS, network-based statistics; RC, resting eyes closed; RO, resting eyes open

**Structural integrity in ABI participants at baseline**

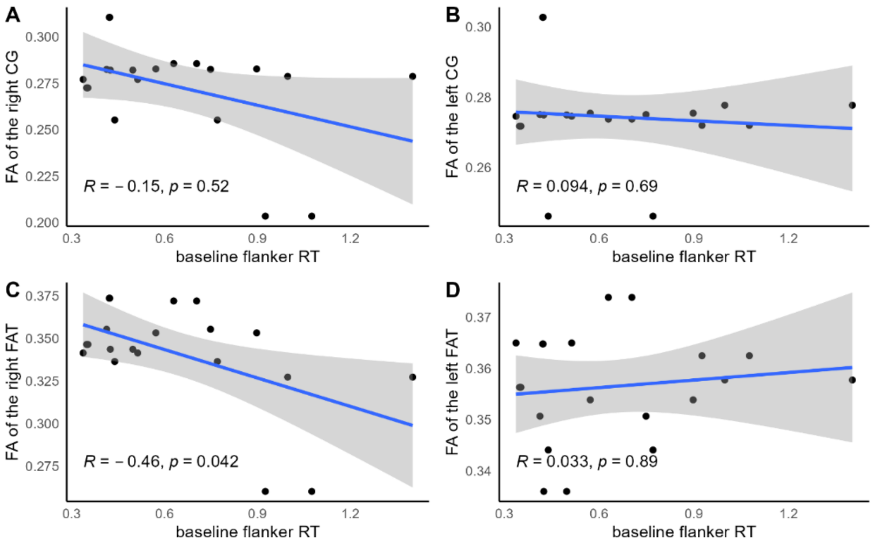


*Supplementary Figure 4: Associations between baseline flanker RT and baseline fractional anisotropy in (A) right CG, (B) left CG, (C) right FAT, (D) left FAT. Greater FA in right FAT associated with faster baseline flanker RT. Spearman's R and 95% CI shown. Baseline data from dlPFC and IFG sessions shown. FA, fractional anisotropy; RT, reaction time; dlPFC, dorsolateral prefrontal cortex; tDCS, transcranial direct current stimulation; FAT, frontal aslant tract; CG, cingulate gyrus; CI, confidence interval.*

#### Association between baseline attention and response to tDCS

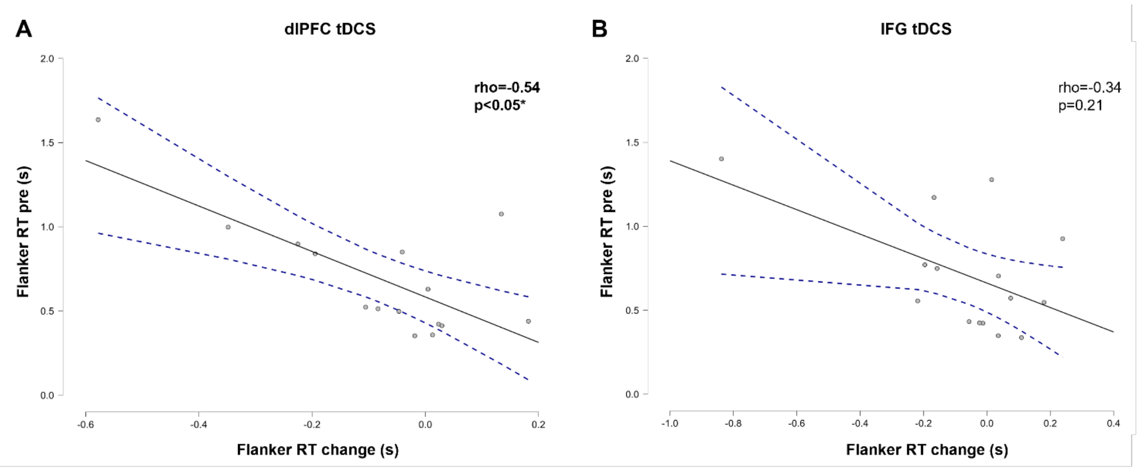


Supplementary Figure 5: Correlations between baseline flanker RT and flanker RT change in ABI participants (A) Slower pre-dlPFC tDCS flanker RT is associated with greater improvement in flanker RT following dlPFC tDCS. (B) Pre-IFG tDCS flanker RT is not associated with flanker RT change following IFG tDCS. Spearman's correlation, 95% confidence interval shown.
RT, reaction time; tDCS, transcranial direct current stimulation; s, seconds; dlPFC, dorsolateral prefrontal cortex; IFG, inferior frontal gyrus

#### Associations between simulated E-field and response to tDCS

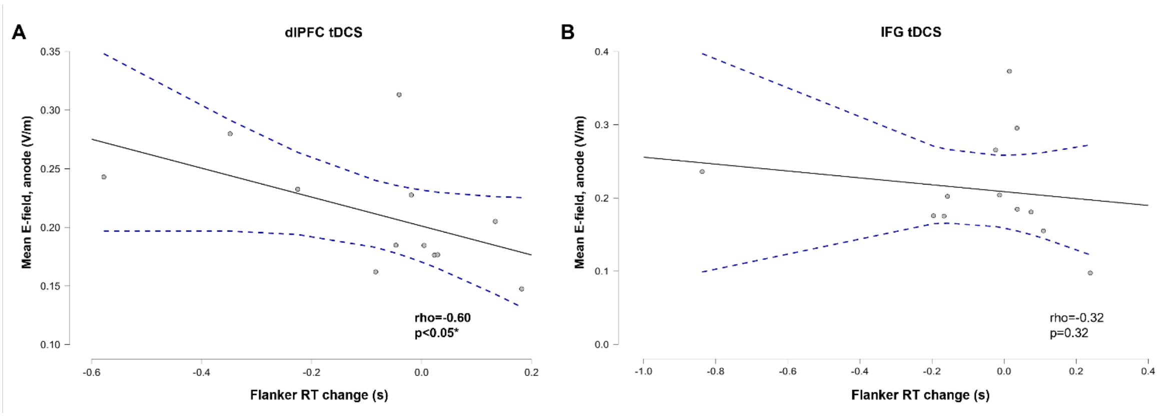


Supplementary Figure 6: Correlations between mean normalised E-field and flanker RT change in ABI participants. (A) Higher E-field is associated with faster flanker RT following dlPFC tDCS. (B) E-field is not associated with flanker RT change following IFG tDCS. Spearman's correlation, 10 mm spherical ROI, 95% confidence interval shown.
dlPFC, dorsolateral prefrontal cortex; E-field, electric field; tDCS, transcranial direct current stimulation; RT, reaction time;

#### Supplementary References


1. Teasdale G, Jennett B, Teasdale G, Jennett B. Glasgow Coma Scale (GCS). Retrieved October 1974;2(7872):81-4.
2. Brooks BL, Sherman EM. Computerized neuropsychological testing to rapidly evaluate cognition in pediatric patients with neurologic disorders. Journal of Child Neurology 2012;27(8):982-91.
3. Gioia GA, Isquith PK, Guy SC, Kenworthy L. BRIEF-2: Behavior rating inventory of executive function. Psychological Assessment Resources Lutz, FL; 2015.
4. McCarthy ML, MacKenzie EJ, Durbin DR, Aitken ME, Jaffe KM, Paidas CN, et al. The Pediatric Quality of Life Inventory: an evaluation of its reliability and validity for children with traumatic brain injury. Archives of Physical Medicine and Rehabilitation 2005;86(10):1901-9.
5. Eriksen BA, Eriksen CW. Effects of noise letters upon the identification of a target letter in a nonsearch task. Perception & Psychophysics 1974;16(1):143-9.
6. Gershon RC, Wagster MV, Hendrie HC, Fox NA, Cook KF, Nowinski CJ. NIH toolbox for assessment of neurological and behavioral function. Neurology 2013;80(11 Supplement 3):S2-S6.
7. Friehs MA, Dechant M, Vedress S, Frings C, Mandryk RL. Effective gamification of the stop-signal task: two controlled laboratory experiments. JMIR Serious Games 2020;8(3):e17810.
8. Logan GD, Cowan WB, Davis KA. On the ability to inhibit simple and choice reaction time responses: a model and a method. Journal of Experimental Psychology: Human Perception Performance 1984;10(2):276.
9. Schroeder PA, Lohmann J, Ninaus M. Preserved Inhibitory Control Deficits of Overweight Participants in a Gamified Stop-Signal Task: Experimental Study of Validity. JMIR Serious Games 2021;9(1):e25063.
10. Vermeir JF, White MJ, Johnson D, Crombez G, Van Ryckeghem DM. The effects of gamification on computerized cognitive training: systematic review and meta-analysis. JMIR serious games 2020;8(3):e18644.
11. Nand K, Baghaei N, Casey J, Barmada B, Mehdipour F, Liang H-N. Engaging children with educational content via Gamification. Smart Learning Environments 2019;6:1-15.
12. Craven MP, Groom MJ. Computer games for user engagement in Attention Deficit Hyperactivity Disorder (ADHD) monitoring and therapy. In: 2015 International Conference on Interactive Technologies and Games: IEEE; 2015. p. 34-40.
13. Donders FC. On the speed of mental processes. Acta Psychologica 1969;30:412-31.
14. Conners CK, Staff M, Connelly V, Campbell S, MacLean M, Barnes J. Conners' continuous performance Test II (CPT II v. 5). Multi-Health Syst Inc 2000;29:175-96.
15. Gordon B, Caramazza A. Lexical decision for open-and closed-class words: Failure to replicate differential frequency sensitivity. Brain and Language 1982;15(1):143-60.
16. Howell DR, Meehan III WP, Barber Foss KD, Reches A, Weiss M, Myer GD. Reduced dual-task gait speed is associated with visual Go/No-Go brain network activation in children and adolescents with concussion. Brain Injury 2018;32(9):1129-34.
17. Votruba KL, Langenecker SA. Factor structure, construct validity, and age-and education-based normative data for the Parametric Go/No-Go Test. Journal of Clinical and Experimental Neuropsychology 2013;35(2):132-46.
18. Levin HS, Hanten G, Zhang L, Swank PR, Hunter J. Selective impairment of inhibition after TBI in children. Journal of Clinical and Experimental Neuropsychology 2004;26(5):589-97.
19. Fertonani A, Rosini S, Cotelli M, Rossini PM, Miniussi C. Naming facilitation induced by transcranial direct current stimulation. Behavioural brain research 2010;208(2):311-8.
20. Angelini M, Calbi M, Ferrari A, Sbriscia-Fioretti B, Franca M, Gallese V, et al. Proactive control strategies for overt and covert go/nogo tasks: an electrical neuroimaging study. PloS one 2016;11(3):e0152188.
21. Calbi M, Siri F, Heimann K, Barratt D, Gallese V, Kolesnikov A, et al. How context influences the interpretation of facial expressions: a source localization high-density EEG study on the "Kuleshov effect". Scientific reports 2019;9(1):2107.

22. Fraschini M, Demuru M, Crobe A, Marrosu F, Stam CJ, Hillebrand A. The effect of epoch length on estimated EEG functional connectivity and brain network organisation. Journal of neural engineering 2016;13(3):036015.
23. Welch P. The use of fast Fourier transform for the estimation of power spectra: a method based on time averaging over short, modified periodograms. IEEE Transactions on audio and electroacoustics 1967;15(2):70-3.
24. Roig-Herrero A, Planchuelo-Gómez Á, Hernández-García M, de Luis-García R, Fernández-Linsenbarth I, Molina V. Default mode network components and its relationship with anomalous self-experiences in schizophrenia: A rs-fMRI exploratory study. Psychiatry Research: Neuroimaging 2022;324:111495.
25. Pimontel MA, Solomonov N, Oberlin L, Kanellopoulos T, Bress JN, Hoptman MJ, et al. Cortical thickness of the salience network and change in apathy following antidepressant treatment for late-life depression. The American Journal of Geriatric Psychiatry 2021;29(3):241-8.
26. Metzler-Baddeley C, Caeyenberghs K, Foley S, Jones DK. Task complexity and location specific changes of cortical thickness in executive and salience networks after working memory training. Neuroimage 2016;130:48-62.
27. Shen Kk, Welton T, Lyon M, McCorkindale AN, Sutherland GT, Burnham S, et al. Structural core of the executive control network: A high angular resolution diffusion MRI study. Human brain mapping 2020;41(5):1226-36.
28. Tournier J-D, Smith R, Raffelt D, Tabbara R, Dhollander T, Pietsch M, et al. MRtrix3: A fast, flexible and open software framework for medical image processing and visualisation. Neuroimage 2019;202:116137.
29. Veraart J, Novikov DS, Christiaens D, Ades-Aron B, Sijbers J, Fieremans E. Denoising of diffusion MRI using random matrix theory. Neuroimage 2016;142:394-406.
30. Kellner E, Dhital B, Kiselev VG, Reisert M. Gibbs-ringing artifact removal based on local subvoxel-shifts. Magnetic resonance in medicine 2016;76(5):1574-81.
31. Andersson JL, Sotiropoulos SN. An integrated approach to correction for off-resonance effects and subject movement in diffusion MR imaging. Neuroimage 2016;125:1063-78.
32. Tustison NJ, Avants BB, Cook PA, Zheng Y, Egan A, Yushkevich PA, et al. N4ITK: improved N3 bias correction. IEEE transactions on medical imaging 2010;29(6):1310-20.
33. Dhollander T, Mito R, Raffelt D, Connelly A. Improved white matter response function estimation for 3-tissue constrained spherical deconvolution. In: Proc. Intl. Soc. Mag. Reson. Med; 2019.
34. Dhollander T, Connelly A. A novel iterative approach to reap the benefits of multi-tissue CSD from just single-shell (+ b= 0) diffusion MRI data. In: Proc ISMRM; 2016. p. 3010.
35. Jeurissen B, Leemans A, Tournier JD, Jones DK, Sijbers J. Investigating the prevalence of complex fiber configurations in white matter tissue with diffusion magnetic resonance imaging. Human brain mapping 2013;34(11):2747-66.
36. Raffelt D, Dhollander T, Tournier J-D, Tabbara R, Smith RE, Pierre E, et al. Bias field correction and intensity normalisation for quantitative analysis of apparent fibre density. In: Proc. Intl. Soc. Mag. Reson. Med; 2017. p. 3541.
37. Raffelt DA, Tournier J-D, Smith RE, Vaughan DN, Jackson G, Ridgway GR, et al. Investigating white matter fibre density and morphology using fixel-based analysis. Neuroimage 2017;144:58-73.
38. Dhollander T, Clemente A, Singh M, Boonstra F, Civier O, Duque JD, et al. Fixel-based analysis of diffusion MRI: methods, applications, challenges and opportunities. Neuroimage 2021;241:118417.
39. Li LM, Violante IR, Zimmerman K, Leech R, Hampshire A, Patel M, et al. Traumatic axonal injury influences the cognitive effect of non-invasive brain stimulation. Brain 2019;142(10):3280-93.
40. La Corte E, Eldahaby D, Greco E, Aquino D, Bertolini G, Levi V, et al. The frontal aslant tract: a systematic review for neurosurgical applications. Frontiers in Neurology 2021;12:641586.
41. Bubb EJ, Metzler-Baddeley C, Aggleton JP. The cingulum bundle: anatomy, function, and dysfunction. Neuroscience & Biobehavioral Reviews 2018;92:104-27.
42. Singh M, Fuelscher I, He J, Anderson V, Silk TJ, Hyde C. Inter-individual performance differences in the stop-signal task are associated with fibre-specific microstructure of the fronto-basal-ganglia circuit in healthy children. Cortex 2021;142:283-95.
43. Singh M, Skippen P, He J, Thomson P, Fuelscher I, Caeyenberghs K, et al. Longitudinal developmental trajectories of inhibition and white-matter maturation of the fronto-basal-ganglia circuits. Developmental Cognitive Neuroscience 2022;58:101171.

44. Wasserthal J, Neher P, Maier-Hein KH. TractSeg-Fast and accurate white matter tract segmentation. NeuroImage 2018;183:239-53.
